# Heterogeneity of survival outcomes in ypN1 breast cancer after neoadjuvant therapy: The role of residual nodal burden in axillary de-escalation

**DOI:** 10.64898/2026.03.04.26347623

**Authors:** Felipe Andrés Cordero da Luz, Rogério Agenor de Araújo, Lúcio Borges de Araújo, Marcelo José Barbosa Silva

**Affiliations:** Center for Cancer Prevention and Research, Uberlandia Cancer Hospital, Av Amazonas n° 1996, Umuarama, Uberlândia, Minas Gerais, CEP 38.405-302, Brazil; Laboratory of Tumor Biomarkers and Osteoimmunology, Department of Immunology, Institute of Biomedical Sciences, Federal University of Uberlandia, Av Pará n° 1720, Bloco 6T, room 07, Umuarama, Uberlândia, Minas Gerais, CEP 38.405-320, Brazil; Medical Faculty, Federal University of Uberlandia, Av Pará n° 1720, Bloco 2H, Umuarama, Uberlândia, Minas Gerais, CEP 38.400-902, Brazil; Institute of Mathematics and Statistics, Federal University of Uberlândia, Av João Naves de Ávila n° 2121. Bloco 1F, room 1F12, Santa Mônica, Uberlândia, Minas Gerais, CEP 38.408-100, Brazil

**Keywords:** Accelerated Failure Time Model, Breast Neoplasms, Cox Regression Analysis, Lymphatic Metastasis, Neoadjuvant Systemic Therapy, Neoplasm Staging

## Abstract

**Background:** The management of residual axillary disease after neoadjuvant therapy (NAT) remains controversial, as current recommendations often treat ypN1 breast cancer as a homogeneous entity despite potential prognostic heterogeneity. Evidence supporting uniform axillary surgical strategies across different levels of residual nodal burden is limited. We investigated whether survival associations related to axillary surgical evaluation differ according to residual nodal burden in ypN1 disease, using an adjuvant cohort to validate a SEER-based proxy for surgical extent.

**Methods:** Patients with 1–3 positive lymph nodes were identified in the SEER database (2000–2022) and stratified into neoadjuvant (NAT; n=30,560) and adjuvant (AT; n=197,586) cohorts. Axillary surgical evaluation was categorized as limited (2–3 examined nodes) or extensive (≥10 examined nodes). Survival was analyzed using Kaplan–Meier methods and log-logistic accelerated failure-time models, adjusted with inverse probability of treatment weighting.

**Results:** In the ypN1 cohort, limited axillary evaluation was not associated with inferior overall survival among patients with a single residual positive node (IPTW-adjusted HR: 1.15, p=0.134; time ratio [TR]: 0.86, p=0.184). In contrast, limited evaluation was associated with worse survival in patients with two positive nodes (HR: 1.70, 95%CI 1.54–1.87; TR: 0.58, 95%CI 0.53–0.64). The findings were similar when using breast cancer-specific survival as the endpoint.

**Conclusions:** Survival associations related to axillary surgical evaluation after NAT vary according to residual nodal burden. Axillary de-escalation appears feasible in patients with a single residual positive node but cannot be extrapolated to those with multiple residual nodes, underscoring heterogeneity within ypN1 disease.

## 1. INTRODUCTION

Sentinel lymph node biopsy (SLNB) has enabled substantial axillary de-escalation in breast cancer surgery, demonstrating oncological safety comparable to axillary lymph node dissection (ALND) while offering improved quality of life [1]. In the adjuvant (upfront surgery) setting, multiple randomized trials established that completion ALND can be safely omitted in selected patients with limited nodal involvement, provided appropriate local and systemic therapies are administered [2–6]. More recently, the SENOMAC trial further expanded these indications, and current American Society of Clinical Oncology (ASCO) guidelines now support omission of ALND in clinically node-negative patients with limited nodal disease undergoing upfront surgery [7, 8]. Importantly, however, these data were generated exclusively in the adjuvant setting and may not be directly extrapolated to other clinical contexts.

In contrast, axillary management after neoadjuvant systemic therapy (NAT) remains an area of ongoing uncertainty. Patients with residual nodal disease (ypN1) are commonly managed with ALND, despite the absence of randomized evidence demonstrating a survival benefit of uniform surgical escalation in this population [2–6]. Existing studies in the post-NAT setting have largely focused on technical performance metrics of SLNB—such as false-negative rates and mapping strategies—rather than long-term oncologic outcomes [9–13]. Moreover, residual nodal disease after NAT represents a distinct biological scenario, potentially enriched for treatment-resistant disease, raising the possibility that prognostic implications may vary according to the extent of residual nodal burden.

Despite this, ypN1 disease is typically treated as a homogeneous category in clinical decision-making. Whether the survival associations related to axillary surgical extent differ between patients with minimal versus greater residual nodal burden (e.g., one versus two positive nodes) remains insufficiently explored. Addressing this gap is critical to defining the boundaries of safe axillary de-escalation after NAT.

In this population-based study, we examined the association between axillary disease burden—quantified by the number of examined and positive lymph nodes—and overall survival among patients receiving NAT. To ensure methodological robustness, parallel analyses were conducted in a large adjuvant cohort, serving as an internal benchmark to validate our surgical proxy and analytic approach by reproducing established level 1 evidence from the upfront surgery setting. This framework allowed for a more reliable interpretation of survival associations in the less-defined neoadjuvant context.

## 2. MATERIALS

### 2.1 Data Source and Study Design

This retrospective, population-based observational study used data from the Surveillance, Epidemiology, and End Results (SEER) program (17 registries, November 2024 submission), covering diagnoses from 2000 to 2022. The study evaluated survival associations according to axillary disease burden and the extent of axillary surgical evaluation, using the number of examined lymph nodes as a proxy for surgical extent.

Patients were stratified into two cohorts based on treatment sequence: (1) a neoadjuvant therapy (NAT) cohort, including patients with residual nodal disease after systemic therapy (ypN1), and (2) an adjuvant therapy (AT) cohort, consisting of patients who underwent upfront surgery (pN1). The AT cohort was analyzed in parallel as an internal methodological benchmark, allowing assessment of whether the surgical proxy and analytic approach reproduced survival patterns consistent with established evidence from the adjuvant setting before application to the NAT cohort.

### 2.2 Ethics Approval

This study used publicly available, de-identified data. According to Resolution No. 510/2016 of the Brazilian National Health Council (CEP/CONEP), analyses of anonymized secondary databases do not require ethics committee approval or informed consent. The National Institutes of Health (NIH) and the SEER program similarly do not require additional approval for the use of de-identified data.

### 2.3 Selection Criteria

Women diagnosed with invasive breast cancer (ICD-O-3 codes 8500, 8520, 8522, 8523, or 8524) and with 1–3 positive lymph nodes were eligible. Exclusion criteria included age <18 years or unknown; missing cause of death; survival <1 day; multiple primary cancers; occult, Paget, or lysed tumors; bilateral disease; absence of definitive breast surgery; and missing data on molecular subtype or nodal yield.

The primary exposure was the extent of axillary surgical evaluation, categorized as limited (2–3 examined lymph nodes) or extensive (≥10 examined lymph nodes). The cohort selection process is illustrated in Supplementary Figure 1 and detailed in the Supplementary Materials.

### 2.4 Statistical Analysis

Clinical and pathological characteristics were summarized using descriptive statistics. Associations between categorical variables were assessed using χ² tests, with effect sizes estimated by Cramér’s V and Kendall’s τ. The primary endpoint was overall survival (OS). The secondary endpoints was breast cancer-specific survival (BCSS).

Survival was initially evaluated using Kaplan–Meier curves, with log-rank, Tarone–Ware, or Breslow tests selected according to censoring patterns. Multivariable Cox proportional hazards models were fitted to estimate hazard ratios (HRs) and 95% confidence intervals (CIs). The proportional hazards assumption was assessed using Schoenfeld residuals (cox.zph) [14, 15]. When violations were identified for the primary exposure (nodal yield), time-dependent effects were modeled using the tt() function with a log(time) transformation. Other covariates violating the proportional hazards assumption were handled using stratified Cox models.

To provide a complementary time-based interpretation and ensure robustness to proportional hazards violations, parametric accelerated failure time (AFT) models were also fitted [15, 16]. Log-logistic, log-normal, and Weibull distributions were evaluated, with the log-logistic model selected based on Akaike and Bayesian information criteria and Cox–Snell residuals [16]. AFT coefficients were exponentiated to yield time ratios (TRs), where TR <1 indicates a reduction in survival time. Concordant inverse patterns between HRs and TRs were used to support the consistency of findings.

Confounding was addressed using propensity score matching (PSM) and inverse probability of treatment weighting (IPTW). PSM employed fuzzy 1:1 matching with a caliper of 0.0001. IPTW used stabilized weights derived from marginal treatment probabilities [17, 18]. Covariate balance was assessed using standardized mean differences (SMDs) [19], with values <0.2 indicating acceptable balance. Variables with residual imbalance were included in doubly robust survival models [20]. Robust standard errors were applied in all weighted analyses. Weight truncation (maximum weights approximately 10, based on upper percentiles) was explored to enhance numerical stability, with truncated models retained given their near-identical estimates [17, 21].

Survival probabilities at 36, 60, and 120 months were estimated using Monte Carlo simulation incorporating parameter uncertainty. Interaction terms between axillary surgical evaluation and key clinical variables — specifically radiation therapy and residual nodal burden (1 vs. 2 positive nodes) — were tested, retaining interactions with p<0.10. All analyses were conducted using R version 4.4.1 and SPSS version 27.0.

## 3. RESULTS

Since 2018, the SEER database has included a qualitative variable describing the type of axillary surgery performed (e.g., SLNB, ALND). However, internal data review revealed substantial misclassification, with numerous cases coded as “SLNB only” reporting more than 10 or 20 examined lymph nodes, a pattern consistent with complete axillary dissection (data not shown). Given this limited reliability for capturing surgical intent, we did not use this qualitative variable. Instead, consistent with prior literature supporting quantitative nodal yield as a robust surrogate for axillary surgical extent [22–27], patients were classified according to the number of pathologically examined lymph nodes: 1 node, 2–3 nodes, 4–9 nodes, and ≥10 nodes. Baseline characteristics across groups are summarized in Table 1.

**Table 1.**
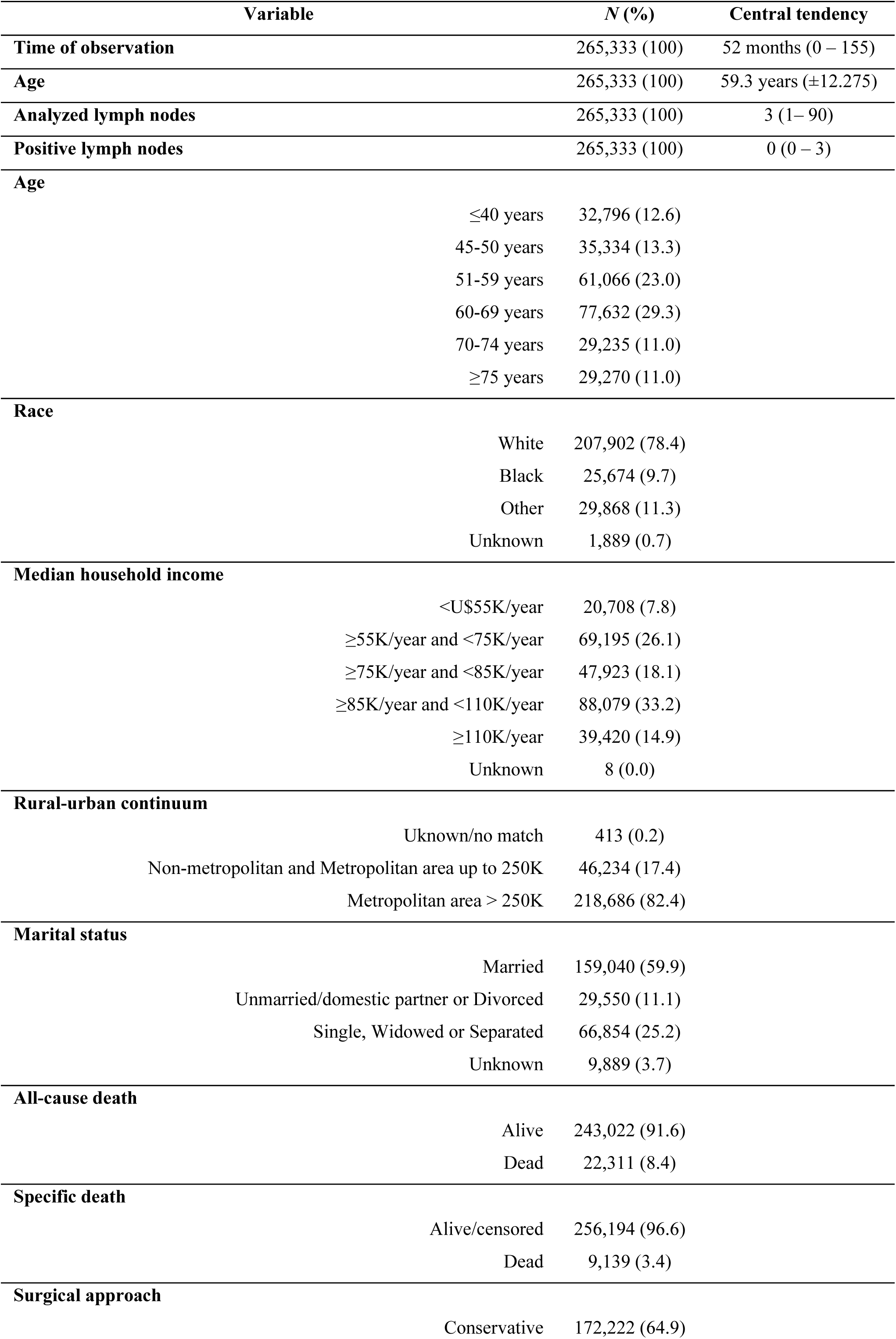
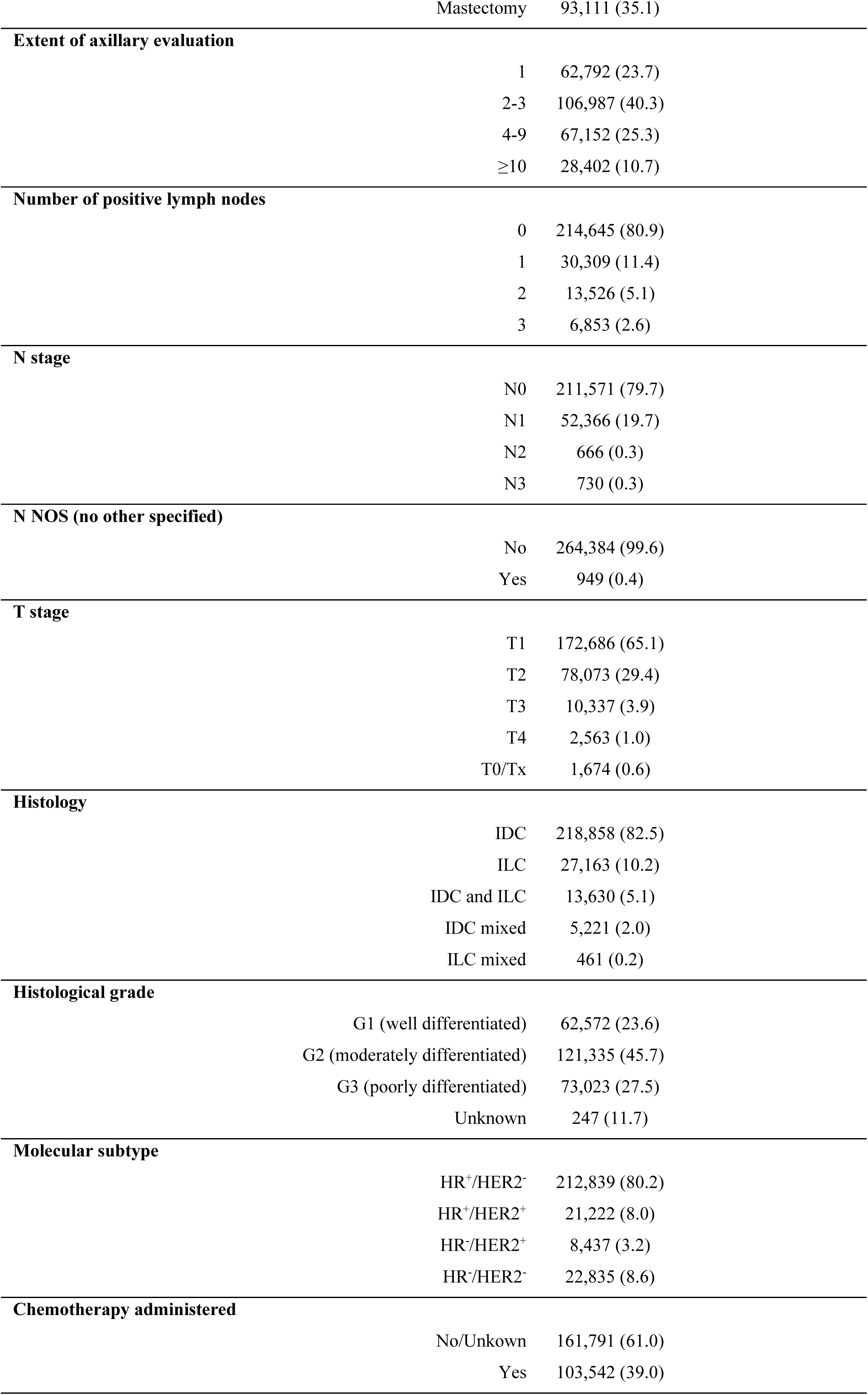
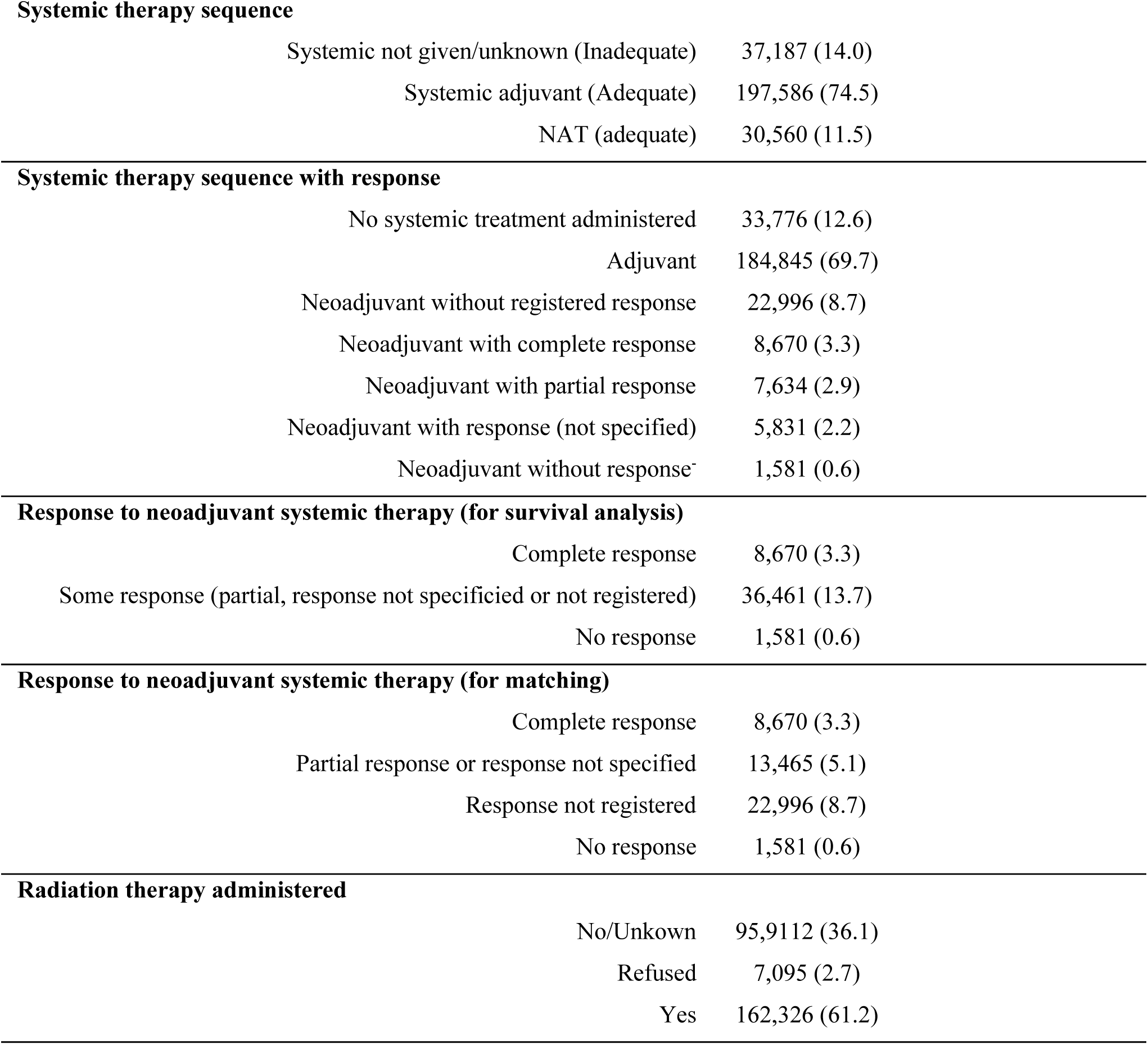
Demographic, clinical, and pathological characteristics (*n*=265,333).

### 3.1 Methodological Validation: The Adjuvant (AT) Cohort

The adjuvant therapy (AT; pN1) cohort was analyzed as an internal methodological benchmark to assess whether the nodal yield–based proxy reproduced survival patterns consistent with established evidence from the upfront surgery setting. Survival curves for these subgroups are shown in Supplementary Figure 2.

Among patients with a single positive lymph node, limited axillary evaluation (2–3 examined nodes) was not associated with inferior overall survival compared with more extensive evaluation (Supplementary Tables 1–3), reproducing survival patterns consistent with those reported in randomized trials supporting axillary de-escalation in this population.

In contrast, among patients with two positive lymph nodes, survival associations differed according to axillary evaluation and regional treatment (Supplementary Tables 4–8). Limited axillary evaluation (2–3 nodes) without radiotherapy was associated with worse survival in both Cox (aHR 1.541, 95%CI 1.154–2.040; p=0.003) and AFT models (aTR 0.676, 95%CI 0.526–0.870; p=0.002) (Supplementary Tables 7). This association was not observed among patients who received radiotherapy (p>0.05, Supplementary Tables 8), suggesting modification of survival associations by regional treatment.

Similar findings were observed for BCSS (Supplementary Tables 9-16; Supplementary Figure 3).

### 3.2 Axillary Evaluation and Survival in the Neoadjuvant (NAT) Cohort

In the neoadjuvant therapy (NAT; ypN1) cohort, survival associations according to axillary evaluation demonstrated a similar dependence on residual nodal burden.

Among patients with a single residual positive node, limited axillary evaluation (2–3 nodes) was not associated with worse overall survival (log-rank p=0.960; IPTW doubly robust-adjusted aHR 0.918, p=0.482), with consistent findings across Cox and AFT models (Table 2; Supplementary Tables 17 and 18).

**Table 2.** Association between the extent of axillary evaluation and overall survival — subgroup of patients with one positive lymph node and treated with neoadjuvant systemic therapy (*n*=5,175).

| Time-dependent Cox regression |  |  |  |  |  |  |  |
| --- | --- | --- | --- | --- | --- | --- | --- |
|  |  | Univariable |  | Multivariable* |  | Multivariable* |  |
| Variable |  | HR (95%CI) | *p* | HR (95%CI) | *p* | HR (95%CI) | *p* |
| Extent of axillary evaluation |  |  |  |  |  |  |  |
|  | ≥10 | 1 |  | 1 |  | 1 |  |
|  | 1 | 1.508 (0.452 – 2.031) | 0.503 | 1.183 (0.917 – 1.526) | 0.195 | 1.183 (0.917 – 1.526) | 0.195 |
|  | 2-3 | 0.563 (0.370 – 0.856) | 0.013 | 0.344 (0.094 – 1.258) | 0.107 | 0.344 (0.094 – 1.258) | 0.107 |
|  | 4-9 | 0.382 (0.147 – 0.990) | 0.048 | 1.023 (0.840 – 1.245) | 0.823 | 1.023 (0.840 – 1.245) | 0.823 |
| tt* analyzed lymph nodes |  |  |  |  |  |  |  |
|  | 1 | 0.924 (0.661 – 1.290) | 0.643 |  |  |  |  |
|  | 2-3 | 1.547 (1.099 – 2.178) | 0.012 | 1.323 (0.928 – 1.886) | 0.122 | 1.323 (0.928 – 1.886) | 0.122 |
|  | 4-9 | 1.287 (0.991 – 1.670) | 0.058 |  |  |  |  |
| Extent of axillary evaluation* Surgery |  |  |  |  |  |  | ----- |
| Extent of axillary evaluation*Radiation therapy |  |  |  |  |  |  | ----- |
| Extent of axillary evaluation*T stage |  |  |  |  |  |  | ----- |
| **Extent of axillary evaluation\*Molecular subtype** |  |  |  |  |  |  | ----- |
| **Log-logistic survival analysis** |  |  |  |  |  |  |  |
|  |  | **Univariable** |  | **Multivariable<sup>#</sup>** |  | **Multivariable<sup>#&</sup>** |  |
| **Variable** |  | **TR (95%CI)** | **p** | **TR (95%CI)** | **p** | **TR (95%CI)** | **p** |
| **Extent of axillary evaluation** |  |  |  |  |  |  |  |
|  | ≥10 | 1 |  | 1 |  | 1 |  |
|  | 1 | 0.881 (0.721 – 1.077) | 0.217 | 0.876 (0.724 – 1.059) | 0.171 | 0.876 (0.724 – 1.059) | 0.171 |
|  | 2-3 | 1.013 (0.837 – 1.226) | 0.894 | 0.969 (0.809 – 1.159) | 0.728 | 0.969 (0.809 – 1.159) | 0.728 |
|  | 4-9 | 1.038 (0.892 – 0.796) | 0.626 | 0.985 (0.854 – 1.135) | 0.831 | 0.985 (0.854 – 1.135) | 0.831 |
| **Extent of axillary evaluation\* Surgery** |  |  |  |  |  |  | ----- |
| **Extent of axillary evaluation\*Radiation therapy** |  |  |  |  |  |  | ----- |
| **Extent of axillary evaluation\*T stage** |  |  |  |  |  |  | ----- |
| **Extent of axillary evaluation\*Molecular subtype** |  |  |  |  |  |  | ----- |

In contrast, among patients with two residual positive nodes, limited axillary evaluation was associated with worse overall survival compared with more extensive evaluation (≥10 nodes) in both Cox (aHR 1.899, 95%CI 1.381–2.612; p<0.0001) and AFT models (aTR 0.587, 95%CI 0.466–0.739; p<0.0001) (Table 3; similar results in matched and IPTW-doubly robust analysis in Supplementary Tables 19 and 20). In IPTW-adjusted analyses, this divergence corresponded to an absolute 10-year survival difference of 19.6% (9.3% – 28.1%). Kaplan–Meier survival curves for patients with one versus two residual positive nodes are presented in Figure 1. Subgroup analyses showed that irradiation does not alter survival in the case of limited axillary evaluation (data not shown).

**Table 3.**
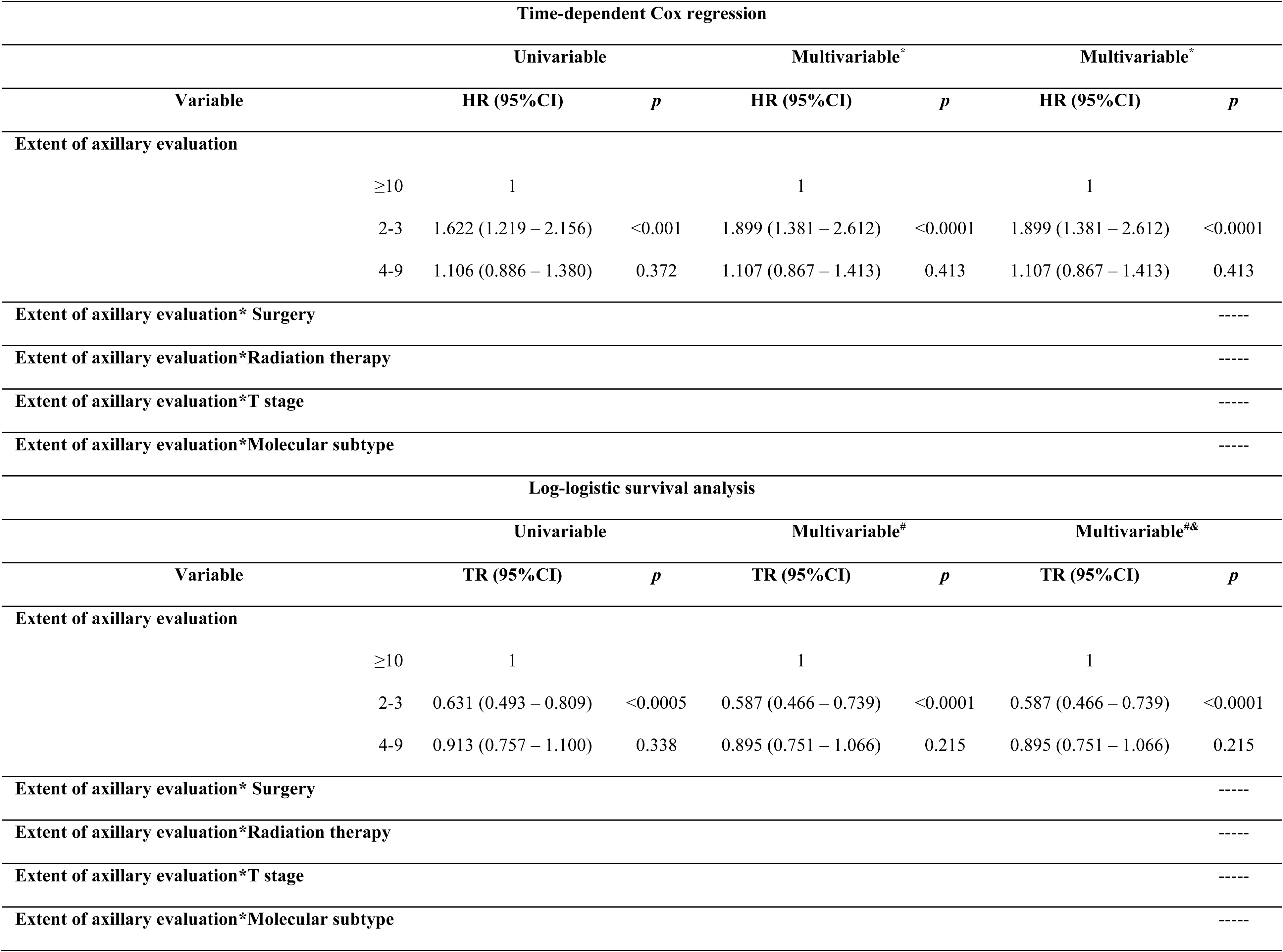
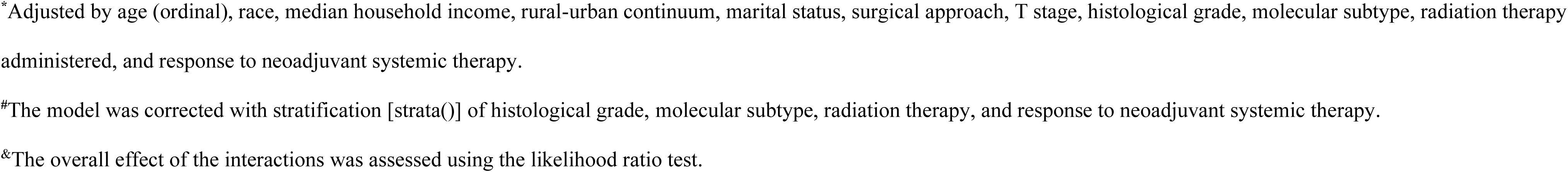
Association between the number of analyzed lymph nodes and overall survival — subgroup of patients with two positive lymph nodes and treated with neoadjuvant systemic therapy (*n*=2,397)

**Figure 1.**
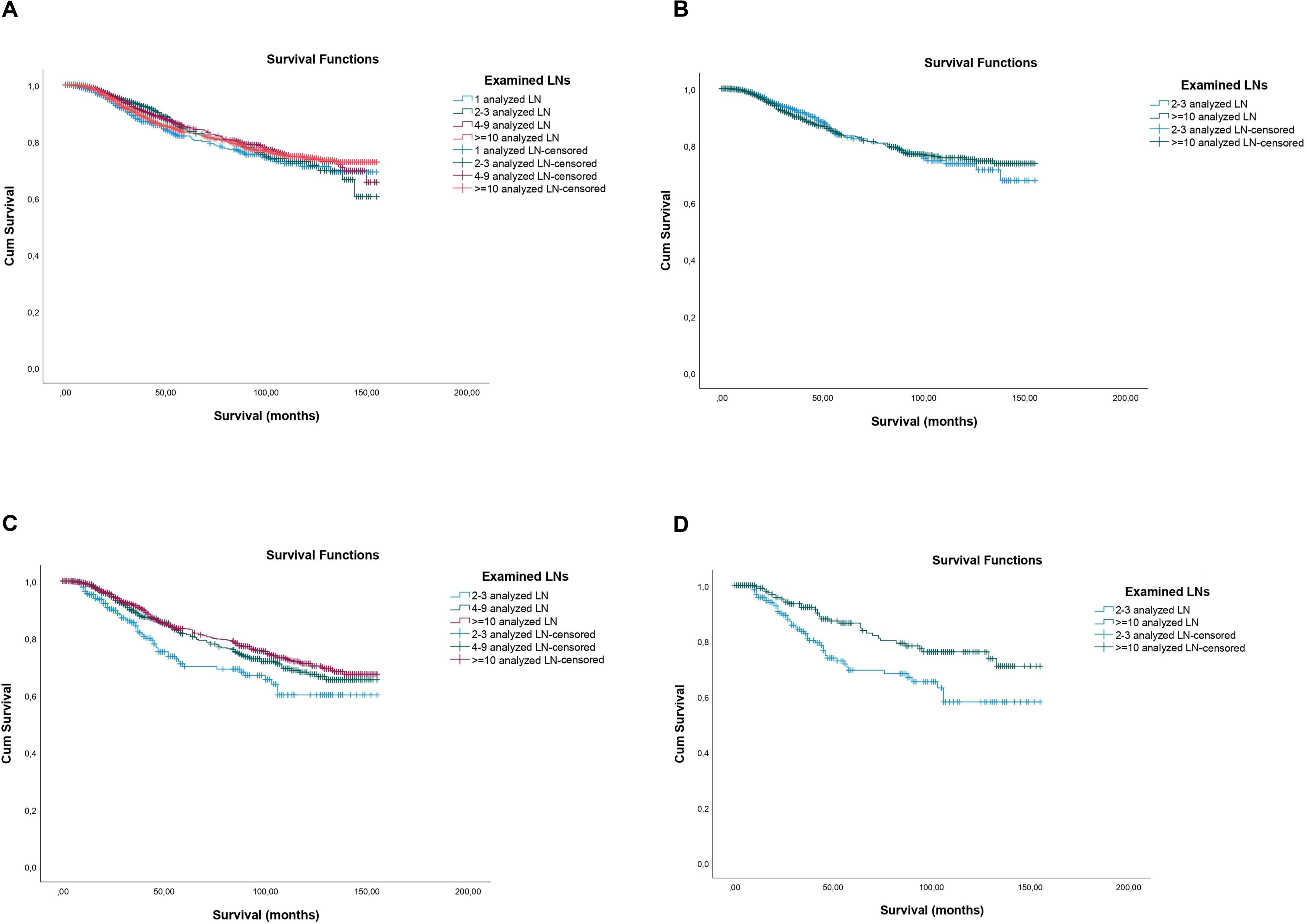
Kaplan-Meier curves for overall survival according to the extent of axillary evaluation in patients submitted to neoadjuvant systemic therapy. (A) Patients with one positive lymph node and treated with neoadjuvant systemic therapy before matching (*n*=5,175) (Log-Rank χ^2^_(df:3)_: 2.37, p=0.500; Breslow: χ^2^_(df:3)_: 6.19, p=0.103; Tarone-Ware: χ^2^_(df:3)_: 4.48; p=0.214). (B) Patients with one positive lymph node and treated with neoadjuvant systemic therapy after matching (*n*=1,482) (Log-Rank χ^2^_(df:3)_: 0.003, p=0.960; Breslow: χ^2^_(df:3)_: 0.27, p=0.601; Tarone-Ware: χ^2^_(df:3)_: 0.09; p=0.766). (C) Patients with two positive lymph node and treated with neoadjuvant systemic therapy before matching (*n*=2,397) (Log-Rank χ^2^_(df:1)_: 11.23, p=0.004; Breslow: χ^2^_(df:1)_: 15.08, p<0.0001; Tarone-Ware: χ^2^_(df:1)_: 13.78; p=0.001. (D) Patients with two positive lymph node and treated with neoadjuvant systemic therapy after matching (*n*=454) (Log-Rank χ^2^_(df:1)_: 8.06, p=0.005; Breslow: χ^2^_(df:1)_: 9.02, p=0.003; Tarone-Ware: χ^2^_(df:1)_: 8.85; p=0.003). Legend: LN – lymph node.

Similar findings were observed for BCSS (Figure 2, Tables 4 and 5, and Supplementary Tables 21 to 24).

**Figure 2.**
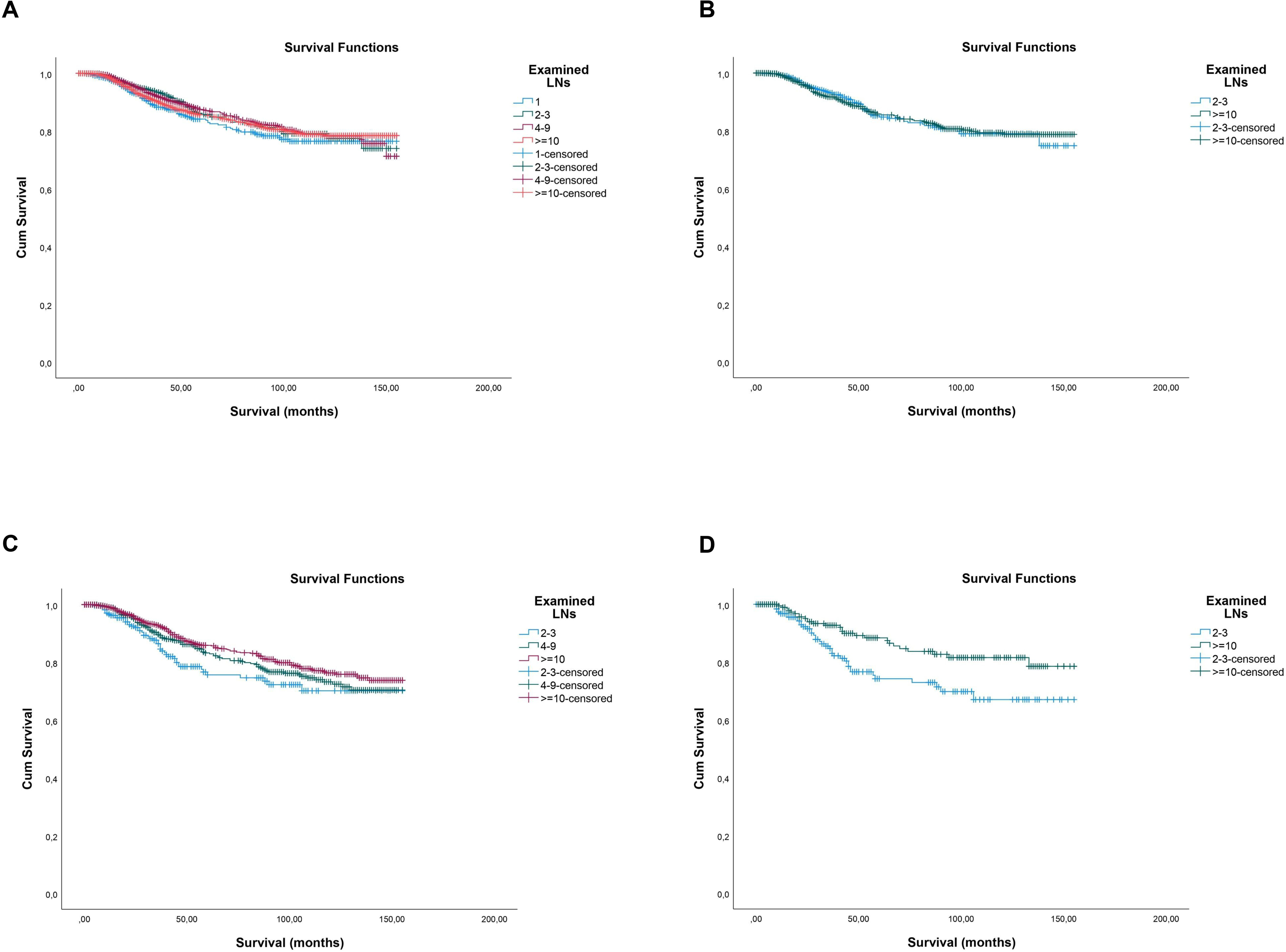
Kaplan-Meier curves for breast cancer-specific survival according to the extent of axillary evaluation in patients submitted to neoadjuvant systemic therapy. (A) Patients with one positive lymph node and treated with neoadjuvant systemic therapy before matching (*n*=5,175) (Log-Rank χ^2^_(df:3)_: 2.86, p=0.413; Breslow: χ^2^_(df:3)_: 7.09, p=0.069; Tarone-Ware: χ^2^_(df:3)_: 5.25; p=0.154). (B) Patients with one positive lymph node and treated with neoadjuvant systemic therapy after matching (*n*=1,482) (Log-Rank χ^2^_(df:3)_: 0.002, p=0.964; Breslow: χ^2^_(df:3)_: 0.17, p=0.676; Tarone-Ware: χ^2^_(df:3)_: 0.07; p=0.794). (C) Patients with two positive lymph node and treated with neoadjuvant systemic therapy before matching (*n*=2,397) (Log-Rank χ^2^_(df:1)_: 7.75, p=0.021; Breslow: χ^2^_(df:1)_: 10.86, p=0.004; Tarone-Ware: χ^2^_(df:1)_: 9.79; p=0.007). (D) Patients with two positive lymph node and treated with neoadjuvant systemic therapy after matching (*n*=454) (Log-Rank χ^2^_(df:1)_: 6.86, p=0.009; Breslow: χ^2^_(df:1)_: 6.97, p=0.008; Tarone-Ware: χ^2^_(df:1)_: 7.15; p=0.007). Legend: LN – lymph node.

**Table 4.**
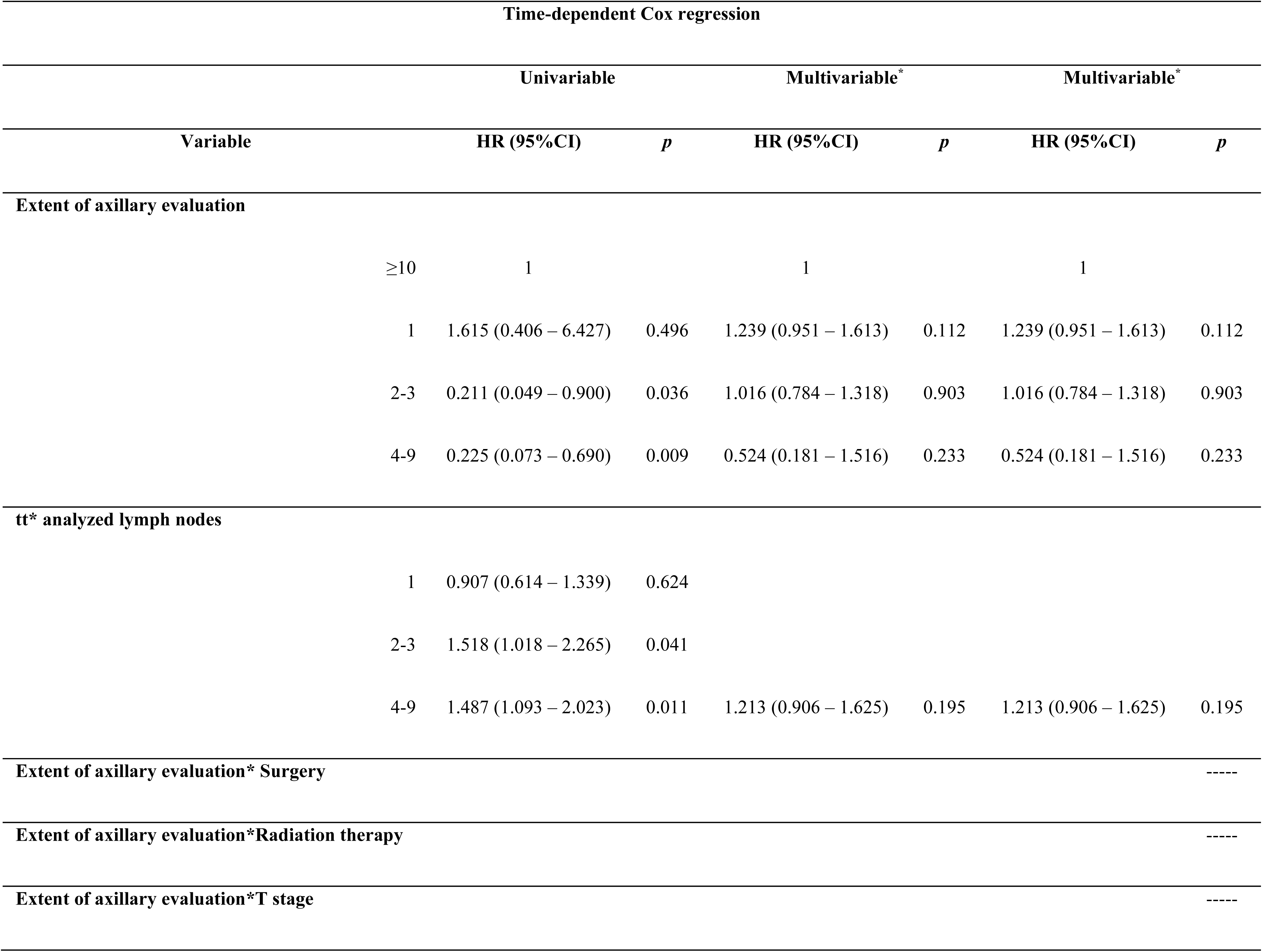
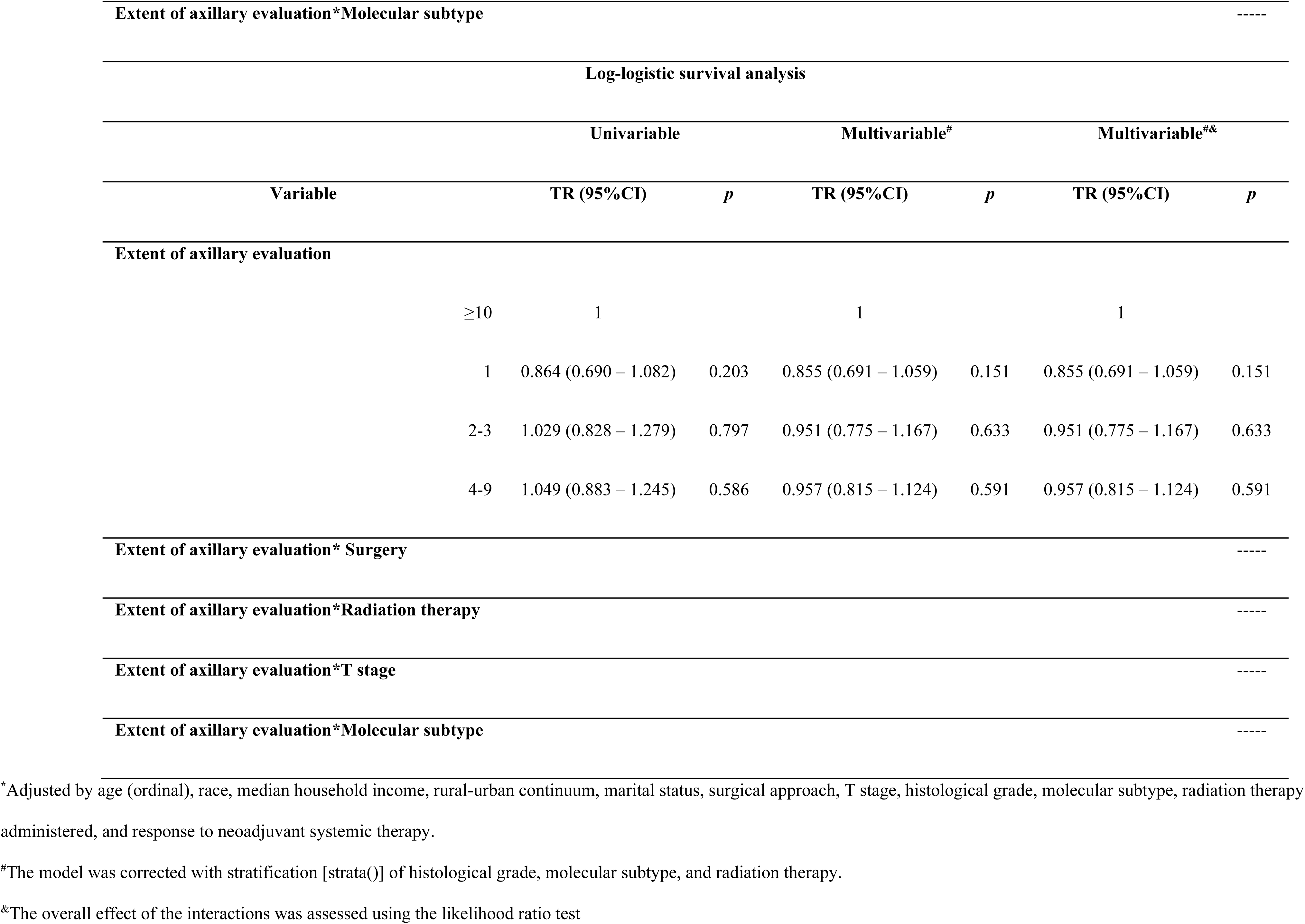
Association between the extent of axillary evaluation and breast cancer-specific survival — subgroup of patients with one positive lymph node and treated with neoadjuvant systemic therapy (*n*=5,175).

**Table 5.**
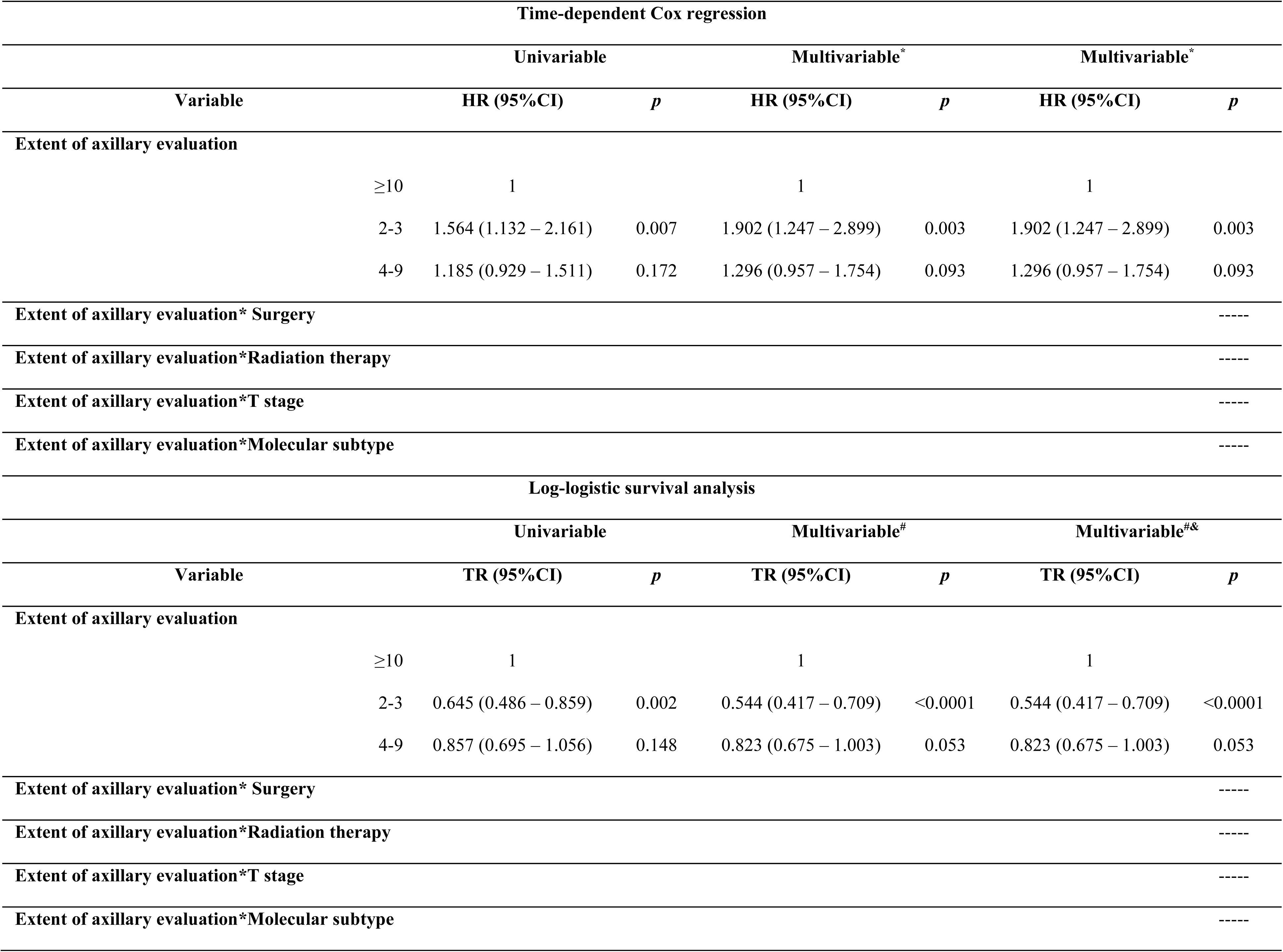
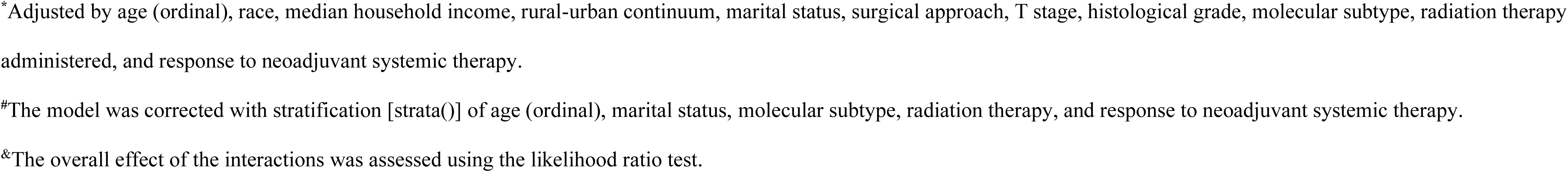
Association between the number of analyzed lymph nodes and breast cancer-specific survival — subgroup of patients with two positive lymph nodes and treated with neoadjuvant systemic therapy (*n*=2,397)

## 4. DISCUSSION

This population-based study provides a large-scale, methodologically validated assessment of survival associations related to axillary surgical evaluation in patients with residual nodal disease after neoadjuvant therapy (NAT). By incorporating an adjuvant cohort as an internal methodological benchmark, we were able to confirm that a nodal yield–based proxy for surgical extent reproduces survival patterns consistent with established level 1 evidence from the upfront surgery setting, including the safety of axillary de-escalation in selected pN1 patients [7, 8]. This validation supports the use of this approach to explore survival associations in the more uncertain post-neoadjuvant context.

The central finding of this study is that survival associations related to axillary evaluation after NAT are not uniform across the ypN1 spectrum. Among patients with a single residual positive node (ypN1=1), limited axillary evaluation (2–3 examined nodes) was not associated with inferior survival, whereas this absence of association was not observed in patients with higher residual nodal burden. In patients with two residual positive nodes (ypN1=2), limited evaluation was consistently associated with worse survival across multiple analytic approaches. Together, these findings suggest that residual nodal burden after NAT identifies biologically and prognostically distinct subgroups, rather than a homogeneous ypN1 population.

This heterogeneity is biologically plausible. Residual nodal disease following NAT may represent selection for tumors with relative treatment resistance and more aggressive behavior, a scenario fundamentally different from treatment-naïve nodal involvement in the adjuvant setting. Our results extend prior literature supporting axillary de-escalation in low-burden nodal disease treated with appropriate radiotherapy [28–32], while demonstrating that such conclusions cannot be assumed to apply uniformly across all levels of residual nodal involvement after NAT.

Our findings also help reconcile previously conflicting observational studies. Almahariq *et al*.[33], reported an overall survival association favoring ALND in ypN1 patients; however, their hazard ratio for patients with a single positive node crossed the null, suggesting uncertainty within that subgroup. By explicitly stratifying ypN1 disease according to residual nodal burden, our analysis shows that survival associations differ meaningfully between ypN1=1 and ypN1=2 patients. Other SEER-based analyses reached divergent conclusions largely due to limited stratification or smaller sample sizes [34–36].

A key methodological strength of this study is the use of formal interaction terms rather than rigid categorical groupings, which can obscure clinically relevant effect modification. Interactions were observed in conditional (matched by propensity score) and marginal-doubly robust (IPTW-doubly robust) models. This approach revealed that regional radiotherapy substantially modifies survival associations related to limited axillary evaluation, particularly in patients with higher nodal burden. Notably, in the adjuvant validation cohort, a higher-risk pattern persisted among mastectomy patients with two positive nodes and minimal nodal evaluation, even when radiotherapy was administered. This observation is consistent with the absence of incidental axillary coverage from tangential breast irradiation in mastectomy patients and reinforces the biological plausibility of our neoadjuvant findings.

Several limitations merit consideration. Surgical extent was inferred using nodal yield as a proxy due to known limitations in SEER coding, locoregional recurrence data were unavailable, and chemotherapy-induced lymphoid depletion may have influenced nodal counts [37]. Nonetheless, the ability of this proxy to reproduce established survival patterns in the adjuvant cohort strongly supports its validity. The use of time-dependent Cox models, complementary accelerated failure time modeling, and robust confounding control further strengthens the reliability of the observed associations.

Importantly, these results should be interpreted as hypothesis-generating and do not replace ongoing prospective randomized trials in the post-neoadjuvant setting, such as Alliance A011202 [38] and TAXIS [39]. Rather, they provide real-world evidence highlighting that uniform axillary strategies may not be appropriate for all ypN1 patients and underscore the importance of risk-adapted approaches while awaiting prospective data.

## 5. CONCLUSION

In this large, validated population-based analysis, survival associations related to axillary surgical evaluation after neoadjuvant therapy varied substantially according to residual nodal burden. Limited axillary evaluation was not associated with inferior survival among patients with a single residual positive node (ypN1=1), whereas similar assumptions of safety could not be extrapolated to patients with higher residual burden (ypN1=2). Regional radiotherapy emerged as an important modifier of these associations, although its ability to fully compensate for minimal axillary evaluation in mastectomy patients warrants further study. While derived from retrospective registry data, these findings highlight clinically relevant heterogeneity within ypN1 disease and support a risk-adapted framework for axillary management in the post-neoadjuvant setting pending results from ongoing randomized trials.

## Authors’ Contribution

F.A.C.L conceived the study and its design, performed data collection, analysis, interpretation and curation, and performed literature search. L.B.A. carried out independent validation of the methodology and results. R.A.A and M.J.B.S. participated in the literature search, data interpretation and supervised the study. All the authors read and approved the final manuscript.

## Data availability

The data that support the findings of this study are available from the National Cancer Institute Surveillance, Epidemiology, and End Results Program, but restrictions apply to the availability of these data, which were used under the license for the current study and so are not publicly available. However, data are available from the authors upon reasonable request and with permission from the National Cancer Institute Surveillance, Epidemiology, and End Results Program. Please contact the corresponding author for any inquiry regarding the data and materials.

