## Supplementary for "Heterogeneity of survival outcomes in ypN1 breast cancer after neoadjuvant therapy: The role of residual nodal burden in axillary de-escalation"

#### Supplementary Materials and Methods

*Selection criteria*

Data extraction was conducted using Boolean filters applied through SEER*Stat software. The following Boolean factor criteria were used to extract data from the SEER database:

{Race, Sex, Year Dx.Sex} = ' Female'

AND {Race and Age (case data only).Age recode with single ages and 90+} != '00 years','01 years','02 years','03 years','04 years','05 years','06 years','07 years','08 years','09 years','10 years','11 years','12 years','13 years','14 years','15 years','16 years','17 years','90+ years','Unknown'

AND {Cause of Death (COD) and Follow-up.SEER other cause of death classification} != 'Dead (missing/unknown COD)','N/A not seq 0-59'

AND {Cause of Death (COD) and Follow-up.SEER cause-specific death classification} != 'Dead (missing/unknown COD)','N/A not seq 0-59'

AND {Cause of Death (COD) and Follow-up.Survival months flag} = 'Complete dates are available and there are more than 0 days of survival'

AND {Multiple Primary Fields.Sequence number} = 'One primary only'

AND {Site and Morphology.Site recode ICD-O-3/WHO 2008} = ' Breast'

AND {Site and Morphology.Behavior code ICD-O-3} = 'Malignant'

AND {Site and Morphology.Histology recode - broad groupings} = '8500-8549: ductal and lobular neoplasms'

AND {Site and Morphology.ICD-O-3 Hist/behav, malignant} = '8500/3: Infiltrating duct carcinoma, NOS','8520/3: Lobular carcinoma, NOS','8522/3: Infiltrating duct and lobular carcinoma','8523/3: Infiltrating duct mixed with other types of carcinoma','8524/3: Infiltrating lobular mixed with other types of carcinoma'

AND {Site and Morphology.Primary Site - labeled} = 'C50.0-Nipple','C50.1-Central portion of breast','C50.2-Upper-inner quadrant of breast','C50.3-Lower-inner quadrant of breast','C50.4-Upper-outer quadrant of breast','C50.5-Lower-outer quadrant of breast','C50.6-Axillary tail of breast','C50.8-Overlapping lesion of breast','C50.9-Breast, NOS'

AND {Site and Morphology.Laterality} = 'Not a paired site','Right - origin of primary','Left - origin of primary','Only one side - side unspecified'

AND {Therapy.RX Summ--Surg Prim Site (1998+)} != 0,9,19,76,90,99

AND {Therapy.Reason no cancer-directed surgery} = 'Surgery performed'

AND {Extent of Disease.Regional nodes positive (1988+)} != 95-99

AND {Extent of Disease.Regional nodes examined (1988+)} != 0,95-99

AND {Extent of Disease.Breast Subtype (2010+)} = 'HR+/HER2+','HR-/HER2+','HR+/HER2-','HR-/HER2-'

AND {Extent of Disease.ER Status Recode Breast Cancer (1990+)} = 'Positive','Negative'

AND {Extent of Disease.PR Status Recode Breast Cancer (1990+)} = 'Positive','Negative'

AND {Extent of Disease.Derived HER2 Recode (2010+)} = 'Positive','Negative'

A total of 427,266 patients' records were retrieved. Subsequently, records form patients with any of the following criteria were excluded: metastatic disease (stage IV); T1 microinvasive disease; *in situ* disease; N1 microinvasive; Nx or lack of N staging; not reported/ inaccurate number (90+) of positive ipsilateral level I-II axillary lymph node(s); presence of positive lymph nodes other than ipsilateral level I-II axillary lymph node(s); systemic therapy surgery sequence other than no systemic therapy, systemic therapy before surgery, surgery both before and after systemic therapy, systemic therapy both before and after surgery, and systemic therapy after surgery; radiation therapy surgery sequence other than no radiation, surgery before radiation, and surgery before and after radiation; radiation therapy other than beam, no/unknown and refused; HR-/HER- or HER2+ disease that did not receive chemotherapy (no/unknown); HR+/HER2- disease with N2/N3 stage that did not receive chemotherapy (no/unknown); time to treatment greater than 4 months; patients undergoing adjuvant systemic therapy, but definitions such as whether they received neoadjuvant therapy in the variable "response to neoadjuvance", and the reverse (received neoadjuvance, but definitions such as "not given" in the variable "response to neoadjuvance).

These exclusion criteria were applied uniformly across all cohorts to ensure internal consistency of comparisons rather than to optimize survival estimates

*Classifications*

As patients with a survival time (follow-up) of less than one day were excluded, all those coded as having 0 months of survival by the system were recoded to 0.03, representing the equivalent of one day of survival.

The staging TNM utilized the seventh edition of the AJCC as the basis for classification. The molecular subtypes were determined by qualitative expression of hormone receptors and HER2 expression.

In the SEER database, the variables ‘Regional Nodes Positive’ and ‘Regional Nodes Examined’ reflect the number of positive and pathologically examined lymph nodes, respectively, with the number of analyzed LNs serving as a surrogate indicator of resected nodes.

For the histological grade, a composite was create using “CS 7 Grade Recode (up to 2017)”, “Grade Clinical (2018+)” and “Grade Pathological (2018+)”. The classification was performed with the CS 7 variable as priority, and the empty values filled with the highest between clinical and pathological grade. Cases classified as 'Undifferentiated/anaplastic; Grade IV' were recoded as grade 3 (poorly differentiated), in alignment with standard practice.

Age was categorized into an ordinal variable with 5-year intervals, grouping patients aged <45 years into a single category; the same was done for patients aged ≥75 years.

For variables with less impact on the study, such as race, histological grade, median household income, marital status and rural-urban continuum code not reported, they were recoded as "Unknown".

The RX Summ--Surg Prim Site (1998+) variable group of categories 20–24 (partial mastectomy) and 30 (subcutaneus mastectomy), 40–49 and 75 (simple mastectomy), 76 (bilateral mastectomy for a single tumor), 50–59 and 63 (modified radical mastectomy), 60–69 and 73–74 (radical mastectomy, NOS), 70–72 (extended radical mastectomy), and 80 (mastectomy, NOS) were recoded as breast-conserving surgery and mastectomy, respectively.

For multicategorical variables, such as age, median household income and others, univariate and multivariate survival analyses were performed to group categories with the same prognostic value regarding overall survival, to reduce the degrees of freedom and facilitate interpretation.

Patients were classified as receiving adjuvant systemic therapy if administrated after surgery. Patients were classified as receiving neoadjuvant therapy (NAT) if systemic therapy was administered before surgery, irrespective of whether additional postoperative systemic therapy was also delivered. These variables were formally used for analysis.

*Statistical analysis*

Descriptive and Bivariate Analyses

Descriptive statistics were computed for all variables. Categorical variables were summarized using absolute and relative frequencies. Continuous variables were assessed for normality via histogram and Q-Q plot inspection. For normally distributed variables, means and standard deviations (SDs) were reported; otherwise, medians with minimum and maximum values were used.

Associations between categorical variables were tested using $\chi^2$ tests. Effect sizes were calculated using Phi and Cramér's V for nominal variables, and Kendall's $\tau$ (b and c) for ordinal variables. For stratified analyses, the Breslow–Day and Tarone tests were used to evaluate homogeneity of odds ratios (ORs), and pooled ORs were calculated using Mantel–Haenszel and Cochran's tests, adjusted for nodal burden.

Survival analysis

Time-to-event analysis was initially performed using Kaplan–Meier (KM) curves. Differences in survival were tested using the log-rank test; when censoring was unbalanced or early/late events predominated, the Tarone–Ware or Breslow test was applied. The proportional hazards (PH) assumption was assessed through Schoenfeld residuals and visual inspection of KM curves.

Multivariable Cox proportional hazards models were used to estimate hazard ratios (HRs) and 95% confidence intervals (CIs). In models adjusted using inverse probability of treatment weighting (IPTW), stabilized and truncated weights were applied, and standard errors were corrected using the robust (sandwich) variance estimator. For models fitted after 1:1 propensity score matching (PSM), standard errors were corrected using cluster-robust variance with clustering by matched pair ID.

In cases of PH violation, time-dependent Cox models were used. Specifically, the main exposure variable (extent of axillary evaluation) was modeled with tt() to account for time-varying effects, while categorical covariates that violated the PH assumption were handled with strata(). For models with time-dependent effects, HRs and corresponding 95% CIs were estimated at specific time points (36, 60, and 120 months) using the model's variance–covariance matrix.

Parametric accelerated failure time (AFT) models with log-logistic distribution were also fitted to estimate time ratios (TRs). The AFT models serve as a robust alternative to Cox models under PH violation, providing a time-scale interpretation of the association between axillary evaluation and surviva.

The log-logistic distribution was selected based on Q–Q plots and AIC/BIC comparisons. Uncertainty intervals for survival probabilities were computed using Monte Carlo simulation from a multivariate normal distribution.

Multivariable regression followed a hierarchical three-block structure:

1. Block 1: Univariable models for the number of lymph nodes examined and its interaction with time to detect time-dependence.
2. Block 2: Addition of clinically relevant and statistically significant covariates, with time interactions applied as needed to address PH violations.
3. Block 3: Inclusion of interaction terms between extent of axillary evaluationand other variables (positive LN count, surgical approach, systemic therapy sequence, radiation therapy, and T/N stage).

Bootstrap validation with 1,000 bias-corrected and accelerated (BCa) resamples was applied when feasible.

Parametric survival analysis

In survival analysis, log-logistic models are a subclass of accelerated failure time (AFT) models. Unlike the Cox proportional hazards model, which estimates hazard ratios (HRs) to describe the relative hazard of the event at any given time, AFT models estimate time ratios (TRs).

A time ratio (TR) represents the multiplicative effect of a covariate on the time to event. A TR greater than 1 indicates an association with a longer time to the event, whereas a TR less than 1 indicates an association with a shorter time to the event. In contrast, Cox models focus on the relative hazard at a given time point, where HR > 1 indicates a higher hazard, whereas HR < 1 indicates an association with lower hazard.

To provide a more interpretable measure of clinical relevance, survival probabilities at specific time points (e.g., 120 months) were derived from the AFT models. These were estimated using the survival function of the log-logistic distribution, applying the predicted linear predictors (lp) from the model. Uncertainty intervals (95% confidence intervals) were computed using simulation from a multivariate normal distribution based on the model's estimated coefficients and variance–covariance matrix.

The global contribution of each covariate to the model was assessed using the Likelihood Ratio Test (LRT). A statistically significant difference between the full model and its nested counterpart (excluding the variable of interest) indicated that the variable meaningfully improved model fit.

Propensity score matching

Predictor variables were selected separately according to the sequence of systemic treatment (neoadjuvant or adjuvant) and nodal burden (1 or 2 positive nodes). Logistic regression models included variables representing the ACOSOG Z0011/SENOMAC eligibility criteria to ensure that the Adjuvant cohort could serve as a methodological benchmarking group:

- Adjuvant regimen with 1 positive lymph node: median household income, rural-urban continuum, histological grade, molecular subtype, surgical approach, T stage, N stage, N-NOS, chemotherapy, and radiation therapy.
- Neoadjuvant regimen with 1 positive lymph node: median household income, rural-urban continuum, histological grade, molecular subtype, surgical approach, chemotherapy, and radiation therapy.
- Adjuvant regimen with 2 positive lymph nodes: median household income, rural-urban continuum, histological grade, molecular subtype, surgical approach, T stage, chemotherapy, and radiation therapy.
- Neoadjuvant regimen with 2 positive lymph nodes: median household income, rural-urban continuum, molecular subtype, surgical approach, T stage, chemotherapy, radiation therapy, and response to neoadjuvant systemic therapy.

The strict caliper of 0.0001 was deliberately chosen to maximize covariate balance between matched groups, even at the expense of sample size reduction. Matching covariates were selected based on clinical relevance, absence of multicollinearity, and alignment with ACOSOG Z0011 criteria. Baseline covariate balance was assessed by calculating the standardized mean difference (SMD) before and after matching. An SMD below 0.2 was considered indicative of acceptable balance, consistent with large-scale observational registry standards. Variables remaining with SMD > 0.2 after weighting were included in doubly robust models.

#### Supplementary Results

#### Supplementary Tables

**Supplementary Table 1.** Association between the extent of axillary evaluation and overall survival — subgroup of patients with one positive lymph node and treated with adjuvant systemic therapy (*n*=22,469).

| Time-dependent Cox regression | | | | | | |
|---|---|---|---|---|---|---|
| | **Univariable** | | **Multivariable*** | | **Multivariable*** | |
| **Variable** | **HR (95%CI)** | ***p*** | **HR (95%CI)** | ***p*** | **HR (95%CI)** | ***p*** |
| **Extent of axillary evaluation** | | | | | | |
| ≥10 | 1 | | 1 | | 1 | |
| 1 | 1.250 (0.687 – 2.274) | 0.464 | 1.330 (1.141 – 1.557) | <0.0001 | 1.330 (1.141 – 1.557) | <0.0001 |
| 2-3 | 0.442 (0.248 – 0.787) | 0.006 | 1.206 (0.672 – 2.164) | 0.531 | 1.206 (0.672 – 2.164) | 0.531 |
| 4-9 | 0.662 (0.393 – 1.116 | 0.121 | 1.172 (1.037 – 1.325) | 0.011 | 1.172 (1.037 – 1.325) | 0.011 |
| **tt* analyzed lymph nodes** | | | | | | |
| 1 | 1.009 (0.864 – 1.178) | 0.912 | | | | |
| 2-3 | 1.227 (1.060 – 1.421) | 0.006 | 0.984 (0.847 – 1.142) | 0.829 | 0.984 (0.847 – 1.142) | 0.829 |
| 4-9 | 1.147 (1.006 – 1.308) | 0.040 | | | | |
| **Extent of axillary evaluation* Surgery** |  |  |  |  |  | ----- |
| **Extent of axillary evaluation*Radiation therapy** |  |  |  |  |  | ----- |
| **Extent of axillary evaluation*T stage** |  |  |  |  |  | ----- |
| **Extent of axillary evaluation*Molecular subtype** |  |  |  |  |  | ----- |
| **Log-logistic survival analysis** | | | | | | |
| | **Univariable** | | **Multivariable#** | | **Multivariable#&** | |
| **Variable** | **TR (95%CI)** | ***p*** | **TR (95%CI)** | ***p*** | **TR (95%CI)** | ***p*** |
| **Extent of axillary evaluation** | | | | | | |
| ≥10 | 1 | | 1 | | 1 | |
| 1 | 0.810 (0.736 – 0.891) | <0.0001 | 0.805 | <0.0001 | 0.805 | <0.0001 |

| | | | | | | | |
|---|---|---|---|---|---|---|---|
| | 2-3 | 1.019 (0.937 – 1.109) | 0.655 | 0.930 (0.853 – 1.015) | 0.103 | 0.930 (0.853 – 1.015) | 0.103 |
| | 4-9 | 0.911 (0.844 – 0.984) | 0.017 | 0.896 (0.830 – 0.968) | 0.005 | 0.896 (0.830 – 0.968) | 0.005 |
| **Extent of axillary evaluation* Surgery** |  |  |  |  |  |  | ----- |
| **Extent of axillary evaluation*Radiation therapy** |  |  |  |  |  |  | ----- |
| **Extent of axillary evaluation*T stage** |  |  |  |  |  |  | ----- |
| **Extent of axillary evaluation*Molecular subtype** |  |  |  |  |  |  | ----- |

[*]Adjusted by age (ordinal), race, median household income, rural-urban continuum, marital status, surgical approach, T stage, histological grade, molecular subtype, radiation therapy administered.

[#] The model was corrected with the stratification [strata()] of age (ordinal), median household income, rural-urban continuum, histological grade, T stage, and radiation therapy administered.

[&]The overall effect of the interactions was assessed using the likelihood ratio test

**Supplementary Table 2.** Association between the extent of axillary evaluation and overall survival — subgroup of patients with one positive lymph nodes and treated with adjuvant systemic therapy after matching (*n*=7,214)

| Time-dependent Cox regression | | | | | | |
|---|---|---|---|---|---|---|
| | **Univariable** | | **Multivariable**[*#] | | **Multivariable**[*#] | |
| **Variable** | **HR (95%CI)** | ***p*** | **HR (95%CI)** | ***p*** | **HR (95%CI)** | ***p*** |
| **Extent of axillary evaluation** | | | | | | |
| ≥10 | 1 | | 1 | | 1 | |
| 2-3 | 1.136 (0.979 – 1.318) | 0.093 | 1.146 (0.979 – 1.341) | 0.089 | 1.102 (0.859 – 1.413) | 0.444 |
| **Extent of axillary evaluation* Surgery** |  |  |  |  |  | ----- |
| **Extent of axillary evaluation*Radiation therapy** |  |  |  |  |  | ----- |
| **Extent of axillary evaluation*T stage** | | | | | | <0.0001 |
| 2-3 analyzed LN*T2 | | | | | 0.998 (0.715 – 1.393) | 0.992 |
| 2-3 analyzed LN*T3 | | | | | 1.493 (0.800 – 2.786) | 0.208 |
| 2-3 analyzed LN*T4 | | | | | 5.160 (1.439 – 18.499) | 0.012 |
| 2-3 analyzed LN*Tx/T0 | | | | | indef | indef |
| **Extent of axillary evaluation*Molecular subtype** |  |  |  |  |  | ----- |
| **Log-logistic survival analysis** | | | | | | |
| | **Univariable** | | **Multivariable**[*] | | **Multivariable**[*&] | |
| **Variable** | **TR (95%CI)** | ***p*** | **TR (95%CI)** | ***p*** | **TR (95%CI)** | ***p*** |
| **Extent of axillary evaluation** | | | | | | |
| ≥10 | 1 | | 1 | | 1 | |
| 2-3 | 0.911 (0.815 – 1.018) | 0.098 | 0.940 (0.843 – 1.048) | 0.264 | 0.940 (0.843 – 1.048) | 0.264 |
| **Extent of axillary evaluation* Surgery** |  |  |  |  |  | ----- |

| | |
|---|---|
| **Extent of axillary evaluation*Radiation therapy** | ----- |
| **Extent of axillary evaluation*T stage** | ----- |
| **Extent of axillary evaluation*Molecular subtype** | ----- |

[*#]Adjusted by age (ordinal), race, median household income, rural-urban continuum, marital status, histological grade, molecular subtype, T stage, N, N NOS, chemotherapy, surgical approach and radiotherapy.

[#]The model was corrected with the stratification [strata()] of age (ordinal), rural-urban continuum, histological grade, molecular subtype, chemotherapy and radiotherapy.

[&]The overall effect of the interactions was assessed using the likelihood ratio test.

**Supplementary Table 3.** Association between the extent of axillary evaluation and overall survival, weighted by inverse propensity of treatment — subgroup of patients with one positive lymph nodes and treated with adjuvant systemic therapy (*n*=12,825)

| Time-dependent Cox regression | | | | | | |
|---|---|---|---|---|---|---|
| | **Univariable** | | **Multivariable*#** | | **Multivariable*#** | |
| **Variable** | **HR (95%CI)** | ***p*** | **HR (95%CI)** | ***p*** | **HR (95%CI)** | ***p*** |
| **Extent of axillary evaluation** | | | | | | |
| ≥10 | 1 | | 1 | | 1 | |
| 2-3 | 1.123 (0.984 – 1.281) | 0.08 | 1.060 (0.925 – 1.214) | 0.399 | 0.945 (0.766 – 1.163) | 0.596 |
| **Extent of axillary evaluation*Surgery** |  |  |  |  |  | ----- |
| **Extent of axillary evaluation*Radiation therapy** |  |  |  |  |  | ----- |
| **Extent of axillary evaluation*T stage** | | | | | | |
| 2-3 analyzed LN*T4 stage | | | | | 5.129 (2.115 – 12.879) | <0.0005 |
| **Extent of axillary evaluation*Molecular subtype** |  |  |  |  |  | ----- |
| **Log-logistic survival analysis** | | | | | | |
| | **Univariable** | | **Multivariable*** | | **Multivariable*&** | |
| **Variable** | **TR (95%CI)** | ***p*** | **TR (95%CI)** | ***p*** | **TR (95%CI)** | ***p*** |
| **Extent of axillary evaluation** | | | | | | |
| ≥10 | 1 | | 1 | | 1 | |
| 2-3 | 0.915 (0.842 – 0.993) | 0.034 | 0.951 (0.878 – 1.030) | 0.217 | 1.033 (0.908 – 1.175) | 0.622 |
| **Extent of axillary evaluation* Surgery** |  |  |  |  |  | ----- |
| **Extent of axillary evaluation*Radiation therapy** |  |  |  |  |  | ----- |
| **Extent of axillary evaluation*T stage** |  |  |  |  |  | ----- |

| | | |
|---|---|---|
| 2-3 analyzed LN*T4 stage | 0.315 (0.159 – 0.624) | 0.001 |
| **Extent of axillary evaluation*Molecular subtype** |  | ----- |

[*]Adjusted by age (ordinal), race, rural-urban continuum, marital status, histological grade, molecular subtype, T stage, N, N NOS, chemotherapy, surgical approach and radiotherapy. Median household income was not included due to perfect collinearity.

[#]The model was corrected with the stratification [strata()] of age (ordinal), rural-urban continuum, histological grade, molecular subtype, T stage, surgical approach, chemotherapy and radiotherapy.

[&]The overall effect of the interactions was assessed using the likelihood ratio test.

**Supplementary Table 4.** Association between the number of analyzed lymph nodes and overall survival — subgroup of patients with two positive lymph nodes and treated with adjuvant systemic therapy (*n*=10,116).

| | | | | | | | |
|---|---|---|---|---|---|---|---|
| **Time-dependent Cox regression** | | | | | | | |
| | | **Univariable** | | **Multivariable$^{*\#}$** | | **Multivariable$^{*\#}$** | |
| **Variable** | | **HR (95%CI)** | ***p*** | **HR (95%CI)** | ***p*** | **HR (95%CI)** | ***p*** |
| **Extent of axillary evaluation** | | | | | | | |
| | ≥10 | 1 | | 1 | | 1 | |
| | 2-3 | 1.272 (1.092 – 1.482) | 0.002 | 1.383 (1.172 – 1.632) | <0.0001 | 1.383 (1.172 – 1.632) | <0.0001 |
| | 4-9 | 0.999 (0.877 – 1.139) | 0.016 | 1.027 (0.895 – 1.178) | 0.705 | 1.027 (0.895 – 1.178) | 0.705 |
| **Extent of axillary evaluation* Surgery** |  |  |  |  |  |  | ----- |
| **Extent of axillary evaluation*Radiation therapy** |  |  |  |  |  |  | ----- |
| **Extent of axillary evaluation*T stage** |  |  |  |  |  |  | ----- |
| **Extent of axillary evaluation*Molecular subtype** |  |  |  |  |  |  | ----- |
| **Log-logistic survival analysis** | | | | | | | |
| | | **Univariable** | | **Multivariable$^{*}$** | | **Multivariable$^{*\&}$** | |
| **Variable** | | **TR (95%CI)** | ***p*** | **TR (95%CI)** | ***p*** | **TR (95%CI)** | ***p*** |
| **Extent of axillary evaluation** | | | | | | | |
| | ≥10 | 1 | | 1 | | 1 | |
| | 2-3 | 0.826 (0.733 – 0.931) | 0.002 | 0.816 (0.722 – 0.922) | 0.001 | 0.816 (0.722 – 0.922) | 0.001 |
| | 4-9 | 0.998 (0.900 – 1.106) | 0.965 | 0.954 (0.862 – 1.055) | 0.360 | 0.954 (0.862 – 1.055) | 0.360 |
| **Extent of axillary evaluation* Surgery** |  |  |  |  |  |  | ----- |
| **Extent of axillary evaluation*Radiation therapy** |  |  |  |  |  |  | ----- |
| **Extent of axillary evaluation*T stage** |  |  |  |  |  |  | ----- |
| **Extent of axillary evaluation*Molecular subtype** |  |  |  |  |  |  | ----- |

[*]Adjusted by age (ordinal), race, median household income, rural-urban continuum, marital status, surgical approach, T stage, histological grade, molecular subtype, radiation therapy administered.

[#]The model was corrected with stratification [strata()] of age (ordinal), histological grade, molecular subtype, and radiation therapy.

[&]The overall effect of the interactions was assessed using the likelihood ratio test

**Supplementary Table 5.** Association between the extent of axillary evaluation and overall survival — subgroup of patients with two positive lymph nodes and treated with adjuvant systemic therapy after matching (*n*=2,820)

| **Time-dependent Cox regression** | | | | | | |
|---|---|---|---|---|---|---|
| | **Univariable** | | **Multivariable*#** | | **Multivariable*#** | |
| **Variable** | **HR (95%CI)** | ***p*** | **HR (95%CI)** | ***p*** | **HR (95%CI)** | ***p*** |
| **Extent of axillary evaluation** | 1 | | 1 | | 1 | |
| ≥10 | 1 | | 1 | | 1 | |
| 2-3 | 1.377 (1.108 – 1.711) | 0.004 | 1.422 (1.128 – 1.793) | 0.003 | 2.227 (1.438 – 3.451) | <0.0005 |
| **Extent of axillary evaluation* Surgery** |  |  |  |  |  | ----- |
| **Extent of axillary evaluation*Radiation therapy** | | | | | | 0.007 |
| 2-3 analyzed LN*received radiation | | | | | 0.530 (0.320 – 0.877) | 0.001 |
| **Extent of axillary evaluation*T stage** |  |  |  |  |  | ----- |
| **Extent of axillary evaluation*Molecular subtype** |  |  |  |  |  | ----- |
| **Log-logistic survival analysis** | | | | | | |
| | **Univariable** | | **Multivariable*** | | **Multivariable*&** | |
| **Variable** | **TR (95%CI)** | ***p*** | **TR (95%CI)** | ***p*** | **TR (95%CI)** | ***p*** |
| **Extent of axillary evaluation** | | | | | | |
| ≥10 | 1 | | 1 | | 1 | |
| 2-3 | 0.796 (0.681 – 0.931) | 0.004 | 0.814 (0.700 – 0.947) | 0.008 | 0.584 (0.435 – 0.783) | 0.0003 |
| **Extent of axillary evaluation* Surgery** |  |  |  |  |  | ----- |
| **Extent of axillary evaluation*Radiation therapy** | | | | | | |
| 2-3 analyzed LN*received radiation | | | | | 1.556 (1.101 – 2.199) | 0.012 |
| **Extent of axillary evaluation*T stage** |  |  |  |  |  | ----- |
| **Extent of axillary evaluation*Molecular subtype** |  |  |  |  |  | ----- |

[*]Adjusted by age (ordinal), race, median household income, rural-urban continuum, marital status, histological grade, molecular subtype, T stage, chemotherapy, surgical approach and radiotherapy.

[#]The model was corrected with stratification [strata()] of histological grade, molecular subtype, chemotherapy, and radiotherapy.

[&]The overall effect of the interactions was assessed using the likelihood ratio test.

**Supplementary Table 6.** Association between the extent of axillary evaluation and overall survival, weighted by inverse propensity of treatment — subgroup of patients with two positive lymph nodes and treated with adjuvant systemic therapy (*n*=6,963)

| | | | | | | | |
|---|---|---|---|---|---|---|---|
| **Time-dependent Cox regression** | | | | | | | |
| | | **Univariable** | | **Multivariable*** | | **Multivariable*#** | |
| **Variable** | | **HR (95%CI)** | ***p*** | **HR (95%CI)** | ***p*** | **HR (95%CI)** | ***p*** |
| **Extent of axillary evaluation** | | | | | | | |
| | ≥10 | 1 | | 1 | | 1 | |
| | 2-3 | 1.334 (1.117 – 1.594) | 0.002 | 1.448 (1.214 – 1.727) | <0.0001 | 1.448 (1.214 – 1.727) | <0.0001 |
| **Extent of axillary evaluation* Surgery** |  |  |  |  |  |  | ----- |
| **Extent of axillary evaluation*Radiation therapy** |  |  |  |  |  |  | ----- |
| **Extent of axillary evaluation*Molecular subtype** |  |  |  |  |  |  | ----- |
| **Log-logistic survival analysis** | | | | | | | |
| | | **Univariable** | | **Multivariable*** | | **Multivariable*&** | |
| **Variable** | | **TR (95%CI)** | ***p*** | **TR (95%CI)** | ***p*** | **TR (95%CI)** | ***p*** |
| **Extent of axillary evaluation** | | | | | | | |
| | ≥10 | 1 | | 1 | | 1 | |
| | 2-3 | 0.794 (0.709 – 0.889) | <0.0001 | 0.798 (0.715 – 0.890) | <0.0001 | 0.664 (0.559 – 0.788) | <0.0001 |
| **Extent of axillary evaluation* Surgery** |  |  |  |  |  |  | ----- |
| **Extent of axillary evaluation*Radiation therapy** | | | | | | | |
| | 2-3 analyzed LN* received radiation | | | | | 1.391 (1.108 – 1.747) | 0.004 |
| **Extent of axillary evaluation*Molecular subtype** |  |  |  |  |  |  | ----- |

*Adjusted by age (ordinal), race, rural-urban continuum, marital status, histological grade, molecular subtype, T stage, chemotherapy, surgical approach and radiotherapy. Median household income was not included due to perfect collinearity.

#The model was corrected with stratification [strata()] of age (ordinal), histological grade, molecular subtype, chemotherapy, and radiotherapy.

[&]The overall effect of the interactions was assessed using the likelihood ratio test.

**Supplementary Table 7.** Association between the extent of axillary evaluation and overall survival — subgroup of patients with two positive lymph nodes, treated with adjuvant systemic therapy and radiation therapy not given/unknown if given (*n*=2,745).

| | | | | | | | |
|---|---|---|---|---|---|---|---|
| **Time-dependent Cox regression** | | | | | | | |
| | | **Univariable** | | **Multivariable*#** | | **Multivariable*#** | |
| **Variable** | | **HR (95%CI)** | ***p*** | **HR (95%CI)** | ***p*** | **HR (95%CI)** | ***p*** |
| **Extent of axillary evaluation** | | | | | | | |
| | ≥10 | 1 | | 1 | | 1 | |
| | 2-3 | 1.786 (1.385 – 2.302) | <0.0001 | 1.541 (1.154 – 2.040) | 0.003 | 1.541 (1.154 – 2.040) | 0.003 |
| | 4-9 | 1.326 (1.073 – 1.638) | 0.009 | 1.244 (1.000 – 1.548) | 0.050 | 1.244 (1.000 – 1.548) | 0.050 |
| **Extent of axillary evaluation* Surgery** |  |  |  |  |  |  | ----- |
| **Extent of axillary evaluation*T stage** |  |  |  |  |  |  | ----- |
| **Log-logistic survival analysis** | | | | | | | |
| | | **Univariable** | | **Multivariable*** | | **Multivariable*&** | |
| **Variable** | | **TR (95%CI)** | ***p*** | **TR (95%CI)** | ***p*** | **TR (95%CI)** | ***p*** |
| **Extent of axillary evaluation** | | | | | | | |
| | ≥10 | 1 | | 1 | | 1 | |
| | 2-3 | 0.555 (0.430 – 0.717) | <0.0001 | 0.676 (0.526 – 0.870) | 0.002 | 0.676 (0.526 – 0.870) | 0.002 |
| | 4-9 | 0.752 (0.611 – 0.925) | 0.007 | 0.822 (0.676 – 0.999) | 0.048 | 0.822 (0.676 – 0.999) | 0.048 |
| **Extent of axillary evaluation* Surgery** |  |  |  |  |  |  | ----- |
| **Extent of axillary evaluation*T stage** |  |  |  |  |  |  | ----- |

*Adjusted by age (ordinal), race, median household income, rural-urban continuum, marital status, surgical approach, T stage, histological grade, and molecular subtype.

#The model was corrected with stratification [strata()] of age (ordinal), molecular subtype, and surgical approach.

[&]The overall effect of the interactions was assessed using the likelihood ratio test

**Supplementary Table 8.** Association between the extent of axillary evaluation and overall survival — subgroup of patients with two positive lymph nodes, treated with adjuvant systemic therapy and radiation therapy (*n*=7,124).

| Time-dependent Cox regression | | | | | | |
|---|---|---|---|---|---|---|
| | Univariable | | Multivariable*# | | Multivariable*# | |
| Variable | HR (95%CI) | *p* | HR (95%CI) | *p* | HR (95%CI) | *p* |
| **Extent of axillary evaluation** | | | | | | |
| ≥10 | 1 | | 1 | | 1 | |
| 2-3 | 1.140 (0.935 – 1.390) | 0.194 | 0.590 (0.185 – 1.885) | 0.373 | 0.590 (0.185 – 1.885) | 0.373 |
| 4-9 | 0.839 (0.704 – 1.000) | 0.050 | 0.896 (0.746 – 1.078) | 0.245 | 0.896 (0.746 – 1.078) | 0.245 |
| **tt* analyzed lymph nodes** | | | | | | |
| 2-3 | | | 1.200 (0.984 – 1.612) | 0.223 | 1.200 (0.984 – 1.612) | 0.223 |
| 4-9 | | | | | | |
| **Extent of axillary evaluation* Surgery** |  |  |  |  |  | ----- |
| **Extent of axillary evaluation*T stage** |  |  |  |  |  | ----- |
| **Log-logistic survival analysis** | | | | | | |
| | Univariable | | Multivariable* | | Multivariable*& | |
| Variable | TR (95%CI) | *p* | TR (95%CI) | *p* | TR (95%CI) | *p* |
| **Extent of axillary evaluation** | | | | 0.096 | | 0.096 |
| ≥10 | 1 | | 1 | | 1 | |
| 2-3 | 0.911 (0.796 – 1.044) | 0.180 | 0.919 (0.801 – 1.055) | 0.231 | 0.919 (0.801 – 1.055) | 0.231 |
| 4-9 | 1.131 (1.002 – 1.276) | 0.046 | 1.083 (0.960 – 1.221) | 0.194 | 1.083 (0.960 – 1.221) | 0.194 |
| **Extent of axillary evaluation* Surgery** |  |  |  |  |  | ----- |
| **Extent of axillary evaluation*T stage** |  |  |  |  |  | ----- |

*Adjusted by age (ordinal), race, median household income, rural-urban continuum, marital status, surgical approach, T stage, histological grade, and molecular subtype.

[#]The model was corrected with stratification [strata()] of age (ordinal), rural-urban continuum, histological grade, and molecular subtype.

[&]The overall effect of the interactions was assessed using the likelihood ratio test

**Supplementary Table 9.** Association between the extent of axillary evaluation and breast cancer-specific survival — subgroup of patients with one positive lymph node and treated with adjuvant systemic therapy (*n*=22,469).

| Time-dependent Cox regression | | | | | | | |
|---|---|---|---|---|---|---|---|
| | | **Univariable** | | **Multivariable[*]** | | **Multivariable[*]** | |
| **Variable** | | **HR (95%CI)** | ***p*** | **HR (95%CI)** | ***p*** | **HR (95%CI)** | ***p*** |
| **Extent of axillary evaluation** | | | | | | | |
| | ≥10 | 1 | | 1 | | 1 | |
| | 1 | 0.900 (0.372 – 2.178) | 0.815 | 1.460 (1.208 – 1.753) | <0.0005 | 1.460 (1.208 – 1.753) | <0.0005 |
| | 2-3 | 0.252 (0.105 – 0.606) | 0.002 | 0.753 (0.326 – 1.741) | 0.508 | 0.753 (0.326 – 1.741) | 0.508 |
| | 4-9 | 0.595 (0.286 – 1.241) | 0.166 | 1.188 (1.028 – 1.374) | 0.020 | 1.188 (1.028 – 1.374) | 0.020 |
| **tt* analyzed lymph nodes** | | | | | | | |
| | 1 | 1.074 (0.851 – 1.356) | 0.547 | | | | |
| | 2-3 | 1.357 (1.083 – 1.700) | 0.008 | 1.085 (0.875 – 1.344) | 0.458 | 1.085 (0.875 – 1.344) | 0.458 |
| | 4-9 | 1.172 ( 0.970 – 1.416) | 0.100 | | | | |
| **Extent of axillary evaluation* Surgery** |  |  |  |  |  |  | ----- |
| **Extent of axillary evaluation*Radiation therapy** |  |  |  |  |  |  | ----- |
| **Extent of axillary evaluation*T stage** |  |  |  |  |  |  | ----- |
| **Extent of axillary evaluation*Molecular subtype** |  |  |  |  |  |  | ----- |
| **Log-logistic survival analysis** | | | | | | | |
| | | **Univariable** | | **Multivariable[#]** | | **Multivariable[#&]** | |
| **Variable** | | **TR (95%CI)** | ***p*** | **TR (95%CI)** | ***p*** | **TR (95%CI)** | ***p*** |
| **Extent of axillary evaluation** | | | | | | | |

| | | | | | | |
|---|---|---|---|---|---|---|
| ≥10 | 1 | | 1 | | 1 | |
| 1 | 0.848 (0.740 – 0.972) | 0.018 | 0.742 (0.644 – 0.854) | <0.0001 | 0.742 (0.644 – 0.854) | <0.0001 |
| 2-3 | 1.157 (1.021 – 0.1312) | 0.023 | 0.949 (0.835 – 1.079) | 0.423 | 0.949 (0.835 – 1.079) | 0.423 |
| 4-9 | 0.917 (0.823 – 1.022) | 0.118 | 0.858 (0.770 – 0.957) | 0.006 | 0.858 (0.770 – 0.957) | 0.006 |
| **Extent of axillary evaluation* Surgery** |  |  |  |  |  | ----- |
| **Extent of axillary evaluation*Radiation therapy** |  |  |  |  |  | ----- |
| **Extent of axillary evaluation*T stage** |  |  |  |  |  | ----- |
| **Extent of axillary evaluation*Molecular subtype** |  |  |  |  |  | ----- |

*Adjusted by age (ordinal), race, median household income, rural-urban continuum, marital status, surgical approach, T stage, histological grade, molecular subtype, radiation therapy administered.

### The model was corrected with the stratification [strata()] of race, median household income, rural-urban continuum, histological grade, molecular subtype, and radiation therapy administered.

&The overall effect of the interactions was assessed using the likelihood ratio test

**Supplementary Table 10.** Association between the extent of axillary evaluation and breast cancer-specific survival — subgroup of patients with one positive lymph nodes and treated with adjuvant systemic therapy after matching (*n*=7,214)

| | | | | | | |
|---|---|---|---|---|---|---|
| **Time-dependent Cox regression** | | | | | | |
| | **Univariable** | | **Multivariable[*#]** | | **Multivariable[*#]** | |
| **Variable** | **HR (95%CI)** | ***p*** | **HR (95%CI)** | ***p*** | **HR (95%CI)** | ***p*** |
| **Extent of axillary evaluation** | | | | | | |
| ≥10 | 1 | | 1 | | 1 | |
| 2-3 | 1.066 (0.866 – 1.313) | 0.545 | 1.080 (0.871 – 1.340) | 0.484 | 1.111 (0.780 – 1.583) | 0.561 |
| **Extent of axillary evaluation* Surgery** |  |  |  |  |  | ----- |
| **Extent of axillary evaluation*Radiation therapy** |  |  |  |  |  | ----- |
| **Extent of axillary evaluation*T stage** | | | | | | |
| 2-3 analyzed LN*T2 |  |  |  |  | ----- | >0.05 |
| 2-3 analyzed LN*T3 |  |  |  |  | ----- | >0.05 |
| 2-3 analyzed LN*T4 |  |  |  |  | ----- | >0.05 |
| 2-3 analyzed LN*Tx/T0 | | | | | indef | indef |
| **Extent of axillary evaluation*Molecular subtype** |  |  |  |  |  | ----- |
| **Log-logistic survival analysis** | | | | | | |
| | **Univariable** | | **Multivariable[*]** | | **Multivariable[*&]** | |
| **Variable** | **TR (95%CI)** | ***p*** | **TR (95%CI)** | ***p*** | **TR (95%CI)** | ***p*** |
| **Extent of axillary evaluation** | | | | | | |
| ≥10 | 1 | | 1 | | 1 | |
| 2-3 | 0.935 (0.797 – 1.096) | 0.407 | 0.936 (0.800 – 1.095) | 0.407 | 0.936 (0.800 – 1.095) | 0.407 |
| **Extent of axillary evaluation* Surgery** |  |  |  |  |  | ----- |
| **Extent of axillary evaluation*Radiation therapy** |  |  |  |  |  | ----- |

| | |
|---|---|
| **Extent of axillary evaluation*T stage** | ----- |
| **Extent of axillary evaluation*Molecular subtype** | ----- |

[*#]Adjusted by age (ordinal), race, median household income, rural-urban continuum, marital status, histological grade, molecular subtype, T stage, N, N NOS, chemotherapy, surgical approach and radiotherapy.

[#]The model was corrected with the stratification [strata()] of histological grade, molecular subtype, and radiotherapy.

[&]The overall effect of the interactions was assessed using the likelihood ratio test.

**Supplementary Table 11.** Association between the extent of axillary evaluation and breast cancer-specific survival, weighted by inverse propensity of treatment — subgroup of patients with one positive lymph nodes and treated with adjuvant systemic therapy (*n*=12,825)

| Time-dependent Cox regression | | | | | | | |
|---|---|---|---|---|---|---|---|
| | | Univariable | | Multivariable[*#] | | Multivariable[*#] | |
| Variable | | HR (95%CI) | *p* | HR (95%CI) | *p* | HR (95%CI) | *p* |
| **Extent of axillary evaluation** | | | | | | | |
| | ≥10 | 1 | | 1 | | 1 | |
| | 2-3 | 1.055 (0.878 – 1.269) | 0.566 | 1.033 (0.864 – 1.234) | 0.722 | 1.033 (0.864 – 1.234) | 0.722 |
| **Extent of axillary evaluation*Surgery** |  |  |  |  |  |  | ----- |
| **Extent of axillary evaluation*Radiation therapy** |  |  |  |  |  |  | ----- |
| **Extent of axillary evaluation*T stage** |  |  |  |  |  |  | ----- |
| **Extent of axillary evaluation*Molecular subtype** |  |  |  |  |  |  | ----- |
| **Log-logistic survival analysis** | | | | | | | |
| | | Univariable | | Multivariable[*] | | Multivariable[*&] | |
| Variable | | TR (95%CI) | *p* | TR (95%CI) | *p* | TR (95%CI) | *p* |
| **Extent of axillary evaluation** | | | | | | | |
| | ≥10 | 1 | | 1 | | 1 | |
| | 2-3 | 0.939 (0.832 – 1.058) | 0.301 | 0.958 (0.851 – 1.078) | 0.474 | 1.029 (0.827 – 1.281) | 0.797 |
| **Extent of axillary evaluation* Surgery** |  |  |  |  |  |  | ----- |
| **Extent of axillary evaluation*Radiation therapy** |  |  |  |  |  |  | ----- |
| **Extent of axillary evaluation*T stage** |  |  |  |  |  |  | ----- |
| **Extent of axillary evaluation*Molecular subtype** |  |  |  |  |  |  | ----- |

| | | |
|---|---|---|
| 2-3*HR-/HER2+ | 0.464 (0.256 – 0.842) | 0.011 |

*Adjusted by age (ordinal), race, rural-urban continuum, marital status, histological grade, molecular subtype, T stage, N, N NOS, chemotherapy, surgical approach and radiotherapy. Median household income was not included due to perfect collinearity.

#The model was corrected with the stratification [strata()] of histological grade, molecular subtype, surgical approach chemotherapy and radiotherapy.

&The overall effect of the interactions was assessed using the likelihood ratio test.

**Supplementary Table 12.** Association between the number of analyzed lymph nodes and breast cancer-specific survival — subgroup of patients with two positive lymph nodes and treated with adjuvant systemic therapy (*n*=10,116).

| | | | | | | | |
|---|---|---|---|---|---|---|---|
| **Time-dependent Cox regression** | | | | | | | |
| | | **Univariable** | | **Multivariable[*#]** | | **Multivariable[*#]** | |
| **Variable** | | **HR (95%CI)** | ***p*** | **HR (95%CI)** | ***p*** | **HR (95%CI)** | ***p*** |
| **Extent of axillary evaluation** | | | | | | | |
| | ≥10 | 1 | | 1 | | 1 | |
| | 2-3 | 1.151 (0.946 – 1.400) | 0.160 | 1.437 (1.168 – 1.768) | 0.0006 | 1.437 (1.168 – 1.768) | 0.0006 |
| | 4-9 | 0.888 (0.749 – 1.053) | 0.171 | 1.016 (0.854 – 1.209) | 0.860 | 1.016 (0.854 – 1.209) | 0.860 |
| **Extent of axillary evaluation* Surgery** |  |  |  |  |  |  | ----- |
| **Extent of axillary evaluation*Radiation therapy** |  |  |  |  |  |  | ----- |
| **Extent of axillary evaluation*T stage** |  |  |  |  |  |  | ----- |
| **Extent of axillary evaluation*Molecular subtype** |  |  |  |  |  |  | ----- |
| **Log-logistic survival analysis** | | | | | | | |
| | | **Univariable** | | **Multivariable[*]** | | **Multivariable[*&]** | |
| **Variable** | | **TR (95%CI)** | ***p*** | **TR (95%CI)** | ***p*** | **TR (95%CI)** | ***p*** |
| **Extent of axillary evaluation** | | | | | | | |
| | ≥10 | 1 | | 1 | | 1 | |
| | 2-3 | 0.864 (0.734 – 1.016) | 0.076 | 0.760 (0.645 – 0.897) | 0.001 | 0.760 (0.645 – 0.897) | 0.001 |
| | 4-9 | 1.089 (0.945 – 1.254) | 0.240 | 0.993 (0.864 – 1.140) | 0.918 | 0.993 (0.864 – 1.140) | 0.918 |
| **Extent of axillary evaluation* Surgery** |  |  |  |  |  |  | ----- |
| **Extent of axillary evaluation*Radiation therapy** |  |  |  |  |  |  | ----- |
| **Extent of axillary evaluation*T stage** |  |  |  |  |  |  | ----- |
| **Extent of axillary evaluation*Molecular subtype** |  |  |  |  |  |  | ----- |

[*]Adjusted by age (ordinal), race, median household income, rural-urban continuum, marital status, surgical approach, T stage, histological grade, molecular subtype, radiation therapy administered.

[#]The model was corrected with stratification [strata()] of histological grade, molecular subtype, and radiation therapy.

[&]The overall effect of the interactions was assessed using the likelihood ratio test

**Supplementary Table 13.** Association between the extent of axillary evaluation and breast cancer-specific survival — subgroup of patients with two positive lymph nodes and treated with adjuvant systemic therapy after matching (*n*=2,820)

| **Time-dependent Cox regression** | | | | | | | |
|---|---|---|---|---|---|---|---|
| | | **Univariable** | | **Multivariable**[*#] | | **Multivariable**[*#] | |
| **Variable** | | **HR (95%CI)** | ***p*** | **HR (95%CI)** | ***p*** | **HR (95%CI)** | ***p*** |
| **Extent of axillary evaluation** | | 1 | | 1 | | 1 | |
| | ≥10 | 1 | | 1 | | 1 | |
| | 2-3 | 1.387 (1.043 – 1.845) | 0.025 | 1.497 (1.107 – 2.024) | 0.009 | 2.951 (1.669 – 5.219) | 0.0002 |
| **Extent of axillary evaluation* Surgery** |  |  |  |  |  |  | ----- |
| **Extent of axillary evaluation*Radiation therapy** | | | | | | | |
| 2-3 analyzed LN*received radiation | | | | | | 0.394 (0.203 – 0.766) | 0.006 |
| **Extent of axillary evaluation*T stage** |  |  |  |  |  |  | ----- |
| **Extent of axillary evaluation*Molecular subtype** |  |  |  |  |  |  | ----- |

| **Log-logistic survival analysis** | | | | | | | |
|---|---|---|---|---|---|---|---|
| | | **Univariable** | | **Multivariable**[*] | | **Multivariable**[*&] | |
| **Variable** | | **TR (95%CI)** | ***p*** | **TR (95%CI)** | ***p*** | **TR (95%CI)** | ***p*** |
| **Extent of axillary evaluation** | | | | | | | |
| | ≥10 | 1 | | 1 | | 1 | |
| | 2-3 | 0.768 (0.626 – 0.941) | <0.001 | 0.771 (0.631 – 0.942) | 0.011 | 0.476 (0.322 – 0.705) | <0.001 |
| **Extent of axillary evaluation* Surgery** |  |  |  |  |  |  | ----- |
| **Extent of axillary evaluation*Radiation therapy** | | | | | | | |
| 2-3 analyzed LN*received radiation | | | | | | 1.876 (1.180 – 2.981) | 0.008 |
| **Extent of axillary evaluation*T stage** |  |  |  |  |  |  | ----- |
| **Extent of axillary evaluation*Molecular subtype** |  |  |  |  |  |  | ----- |

*Adjusted by age (ordinal), race, median household income, rural-urban continuum, marital status, histological grade, molecular subtype, T stage, chemotherapy, surgical approach and radiotherapy.

#The model was corrected with stratification [strata()] of histological grade, molecular subtype, and radiotherapy.

&The overall effect of the interactions was assessed using the likelihood ratio test.

**Supplementary Table 14.** Association between the extent of axillary evaluation and breast cancer-specific survival, weighted by inverse propensity of treatment — subgroup of patients with two positive lymph nodes and treated with adjuvant systemic therapy (*n*=6,963)

**Time-dependent Cox regression**

| | | Univariable | | Multivariable* | | Multivariable*# | |
|---|---|---|---|---|---|---|---|
| **Variable** | | **HR (95%CI)** | ***p*** | **HR (95%CI)** | ***p*** | **HR (95%CI)** | ***p*** |
| **Extent of axillary evaluation** | | | | | | | |
| | ≥10 | 1 | | 1 | | 1 | |
| | 2-3 | 1.388 (1.107 – 1.741) | 0.004 | 1.468 (1.170 – 1.843) | <0.001 | 2.258 (0.719 – 7.091) | 0.163 |
| **Extent of axillary evaluation* Surgery** |  |  |  |  |  |  | ----- |
| **Extent of axillary evaluation*Radiation therapy** |  |  |  |  |  |  | ----- |
| 2-3 analyzed LN* received radiation | | | | | | 0.589 (0.373 – 0.930) | 0.023 |
| **Extent of axillary evaluation*Molecular subtype** |  |  |  |  |  |  | ----- |

**Log-logistic survival analysis**

| | | Univariable | | Multivariable* | | Multivariable*& | |
|---|---|---|---|---|---|---|---|
| **Variable** | | **TR (95%CI)** | ***p*** | **TR (95%CI)** | ***p*** | **TR (95%CI)** | ***p*** |
| **Extent of axillary evaluation** | | | | | | | |
| | ≥10 | 1 | | 1 | | 1 | |
| | 2-3 | 0.741 (0.638 – 0.862) | <0.0001 | 0.743 (0.641 – 0.862) | <0.0001 | 0.571 (0.456 – 0.714) | <0.0001 |
| **Extent of axillary evaluation* Surgery** |  |  |  |  |  |  | ----- |
| **Extent of axillary evaluation*Radiation therapy** | | | | | | | |
| 2-3 analyzed LN* received radiation | | | | | | 1.633 (1.200 – 2.224) | 0.002 |
| **Extent of axillary evaluation*Molecular subtype** |  |  |  |  |  |  | ----- |

*Adjusted by age (ordinal), race, rural-urban continuum, marital status, histological grade, molecular subtype, T stage, chemotherapy, surgical approach and radiotherapy. Median household income was not included due to perfect collinearity.

[#]The model was corrected with stratification [strata()] of histological grade, molecular subtype, chemotherapy, and radiotherapy.

[&]The overall effect of the interactions was assessed using the likelihood ratio test.

**Supplementary Table 15.** Association between the extent of axillary evaluation and breast cancer-specific survival — subgroup of patients with two positive lymph nodes, treated with adjuvant systemic therapy and radiation therapy not given/unknown if given ($n$=2,745).

| Time-dependent Cox regression | | | | | | | |
|---|---|---|---|---|---|---|---|
| | | Univariable | | Multivariable[*#] | | Multivariable[*#] | |
| Variable | | HR (95%CI) | *p* | HR (95%CI) | *p* | HR (95%CI) | *p* |
| **Extent of axillary evaluation** | | | | | | | |
| | ≥10 | 1 | | 1 | | 1 | |
| | 2-3 | 1.894 (1.386 – 2.590) | <0.0005 | 1.823 (1.272 – 2.615) | 0.001 | 1.823 (1.272 – 2.615) | 0.001 |
| | 4-9 | 0.993 (0.738 – 1.337) | 0.954 | 0.980 (0.720 – 1.333) | 0.896 | 0.980 (0.720 – 1.333) | 0.896 |
| **Extent of axillary evaluation* Surgery** |  |  |  |  |  |  | ----- |
| **Extent of axillary evaluation*T stage** |  |  |  |  |  |  | ----- |
| **Log-logistic survival analysis** | | | | | | | |
| | | Univariable | | Multivariable[*] | | Multivariable[*&] | |
| Variable | | TR (95%CI) | *p* | TR (95%CI) | *p* | TR (95%CI) | *p* |
| **Extent of axillary evaluation** | | | | | | | |
| | ≥10 | 1 | | 1 | | 1 | |
| | 2-3 | 0.503 (0.364 – 0.694) | <0.0001 | 0.561 (0.407 – 0.772) | <0.0001 | 0.561 (0.407 – 0.772) | <0.0001 |
| | 4-9 | 0.985 (0.734 – 1.322) | 0.919 | 1.039 (0.787 – 1.370) | 0.789 | 1.039 (0.787 – 1.370) | 0.789 |
| **Extent of axillary evaluation* Surgery** |  |  |  |  |  |  | ----- |
| **Extent of axillary evaluation*T stage** |  |  |  |  |  |  | ----- |

[*]Adjusted by age (ordinal), race, median household income, rural-urban continuum, marital status, surgical approach, T stage, histological grade, and molecular subtype.

[#]The model was corrected with stratification [strata()] of histological grade, molecular subtype, surgical approach, and T stage.

[&]The overall effect of the interactions was assessed using the likelihood ratio test

**Supplementary Table 16.** Association between the extent of axillary evaluation and breast cancer-specific survival — subgroup of patients with two positive lymph nodes, treated with adjuvant systemic therapy and radiation therapy (*n*=7,124).

| **Time-dependent Cox regression** | | | | | | | |
|---|---|---|---|---|---|---|---|
| | | **Univariable** | | **Multivariable[*#]** | | **Multivariable[*#]** | |
| **Variable** | | **HR (95%CI)** | ***p*** | **HR (95%CI)** | ***p*** | **HR (95%CI)** | ***p*** |
| **Extent of axillary evaluation** | | | | | | | |
| | ≥10 | 1 | | 1 | | 1 | |
| | 2-3 | 0.931 (0.718 – 1.208) | 0.592 | 1.171 (0.892 – 1.537) | 0.255 | 1.171 (0.892 – 1.537) | 0.255 |
| | 4-9 | 0.786 (0.629 – 0.982) | 0.034 | 0.942 (0.752 – 1.182) | 0.607 | 0.942 (0.752 – 1.182) | 0.607 |
| **Extent of axillary evaluation* Surgery** |  |  |  |  |  |  | ----- |
| **Extent of axillary evaluation*T stage** |  |  |  |  |  |  | ----- |
| **Log-logistic survival analysis** | | | | | | | |
| | | **Univariable** | | **Multivariable[*]** | | **Multivariable[*&]** | |
| **Variable** | | **TR (95%CI)** | ***p*** | **TR (95%CI)** | ***p*** | **TR (95%CI)** | ***p*** |
| **Extent of axillary evaluation** | | | | | | | |
| | ≥10 | 1 | | 1 | | 1 | |
| | 2-3 | 1.024 (0.844 – 1.244) | 0.807 | 0.903 (0.743 – 1.099) | 0.309 | 0.903 (0.743 – 1.099) | 0.309 |
| | 4-9 | 1.187 (1.005 – 1.403) | 0.044 | 1.058 (0.897 – 1.249) | 0.500 | 1.058 (0.897 – 1.249) | 0.500 |
| **Extent of axillary evaluation* Surgery** |  |  |  |  |  |  | ----- |
| **Extent of axillary evaluation*T stage** |  |  |  |  |  |  | ----- |

[*]Adjusted by age (ordinal), race, median household income, rural-urban continuum, marital status, surgical approach, T stage, histological grade, and molecular subtype.

[#]The model was corrected with stratification [strata()] of histological grade, and molecular subtype

[&]The overall effect of the interactions was assessed using the likelihood ratio test

**Supplementary Table 17.** Association between the extent of axillary evaluation and overall survival — subgroup of patients with one positive lymph nodes and treated with neoadjuvant systemic therapy after matching ($n$=1,482)

| **Time-dependent Cox regression** | | | | | | | |
|---|---|---|---|---|---|---|---|
| | | **Univariable** | | **Multivariable*#** | | **Multivariable*#** | |
| **Variable** | | **HR (95%CI)** | ***p*** | **HR (95%CI)** | ***p*** | **HR (95%CI)** | ***p*** |
| **Extent of axillary evaluation** | | | | | | | |
| | ≥10 | 1 | | 1 | | 1 | |
| | 2-3 | 1.007 (0.761 – 1.334) | 0.959 | 0.964 (0.716 – 1.297) | 0.808 | 0.964 (0.716 – 1.297) | 0.808 |
| **Extent of axillary evaluation* Surgery** |  |  |  |  |  |  | ----- |
| **Extent of axillary evaluation*Radiation therapy** |  |  |  |  |  |  | ----- |
| **Extent of axillary evaluation*T stage** |  |  |  |  |  |  | ----- |
| **Extent of axillary evaluation*Molecular subtype** |  |  |  |  |  |  | ----- |
| **Log-logistic survival analysis** | | | | | | | |
| | | **Univariable** | | **Multivariable*** | | **Multivariable*&** | |
| **Variable** | | **TR (95%CI)** | ***p*** | **TR (95%CI)** | ***p*** | **TR (95%CI)** | ***p*** |
| **Extent of axillary evaluation** | | | | | | | |
| | ≥10 | 1 | | 1 | | 1 | |
| | 2-3 | 0.962 (0.756 – 1.223) | 0.750 | 1.010 (0.806 – 1.265) | 0.933 | 1.010 (0.806 – 1.265) | 0.933 |
| **Extent of axillary evaluation* Surgery** |  |  |  |  |  |  | ----- |
| **Extent of axillary evaluation*Radiation therapy** |  |  |  |  |  |  | ----- |
| **Extent of axillary evaluation*T stage** |  |  |  |  |  |  | ----- |
| **Extent of axillary evaluation*Molecular subtype** |  |  |  |  |  |  | ----- |

*Adjusted by age (ordinal), race, median household income, rural-urban continuum, marital status, histological grade, molecular subtype, T stage, chemotherapy, surgical approach and radiotherapy.

#The model was corrected with the stratification [strata()] of histological grade.

&The overall effect of the interactions was assessed using the likelihood ratio test

**Supplementary Table 18.** Association between the extent of axillary evaluation and overall survival, weighted by inverse propensity of treatment — subgroup of patients with one positive lymph nodes and treated with neoadjuvant systemic therapy (*n*=2,957)

| Time-dependent Cox regression | | | | | | | |
|---|---|---|---|---|---|---|---|
| | | Univariable | | Multivariable*# | | Multivariable*# | |
| Variable | | HR (95%CI) | *p* | HR (95%CI) | *p* | HR (95%CI) | *p* |
| **Extent of axillary evaluation** | | | | | | | |
| | ≥10 | 1 | | 1 | | 1 | |
| | 2-3 | 1.076 (0.847 – 1.368) | 0.549 | 1.023 (0.807 – 1.296) | 0.853 | 1.023 (0.807 – 1.296) | 0.853 |
| **Extent of axillary evaluation* Surgery** |  |  |  |  |  |  | ----- |
| **Extent of axillary evaluation*Radiation therapy** |  |  |  |  |  |  | ----- |
| **Extent of axillary evaluation*T stage** |  |  |  |  |  |  | ----- |
| **Extent of axillary evaluation*Molecular subtype** |  |  |  |  |  |  | ----- |
| **Log-logistic survival analysis** | | | | | | | |
| | | Univariable | | Multivariable* | | Multivariable*& | |
| Variable | | TR (95%CI) | *p* | TR (95%CI) | *p* | TR (95%CI) | *p* |
| **Extent of axillary evaluation** | | | | | | | |
| | ≥10 | 1 | | 1 | | 1 | |
| | 2-3 | 0.915 (0.760 – 1.102) | 0.351 | 0.984 (0.827 – 1.171) | 0.857 | 0.918 (0.723 – 1.166) | 0.482 |
| **Extent of axillary evaluation* Surgery** |  |  |  |  |  |  | ----- |
| **Extent of axillary evaluation*Radiation therapy** |  |  |  |  |  |  | ----- |
| **Extent of axillary evaluation*T stage** |  |  |  |  |  |  | ----- |
| **Extent of axillary evaluation*Molecular subtype** | | | | | | | |

| | | |
|---|---|---|
| 2-3 analyzed LN*$HR^+/HER2^+$ | 3.240 (1.050 – 10.003) | 0.041 |
| 2-3 analyzed LN*$HR^-/HER2^+$ | 0.741 (0.398 – 1.381) | 0.346 |
| 2-3 analyzed LN*$HR^-/HER2^-$ | 1.118 (0.756 – 1.654) | 0.575 |

[*]Adjusted by age (ordinal), race, rural-urban continuum, marital status, histological grade, molecular subtype, T stage, chemotherapy, surgical approach and radiotherapy. Median household income was not included due to perfect collinearity.

[#]The model was corrected with the stratification [strata()] of histological grade, molecular subtype, and chemotherapy.

[&]The overall effect of the interactions was assessed using the likelihood ratio test.

**Supplementary Table 19.** Association between the extent of axillary evaluation and breast overall survival — subgroup of patients with two positive lymph nodes and treated with neoadjuvant systemic therapy after matching (*n*=454)

| Cox regression | | | | | | |
|---|---|---|---|---|---|---|
| | Univariable | | Multivariable* | | Multivariable* | |
| **Variable** | **HR (95%CI)** | ***p*** | **HR (95%CI)** | ***p*** | **HR (95%CI)** | ***p*** |
| **Extent of axillary evaluation** | | | | | | |
| ≥10 | 1 | | 1 | | 1 | |
| 2-3 | 1.872 (1.236 – 2.835) | 0.003 | 2.449 (1.499 – 4.002) | <0.0005 | 1.512 (0.233 – 9.084) | 0.665 |
| **Extent of axillary evaluation* Surgery** |  |  |  |  |  | ----- |
| **Extent of axillary evaluation*Radiation therapy** | | | | | | |
| 2-3 analyzed LN*received radiation | | | | | 0.266 (0.091 – 0.779) | 0.016 |
| **Extent of axillary evaluation*T stage** | | | | | | 0.027 |
| 2-3 analyzed LN*T2 | | | | | 6.930 (1.063 – 45.154) | 0.043 |
| 2-3 analyzed LN*T3 | | | | | 6.097 (0.912 – 40.774) | 0.062 |
| 2-3 analyzed LN*T4 | | | | | 5.098 (0.707 – 36.782) | 0.106 |
| 2-3 analyzed LN*Tx/T0 | | | | | 0.716 (0.065 – 7.891) | 0.785 |
| **Extent of axillary evaluation*Molecular subtype** |  |  |  |  |  | ----- |
| **Log-logistic survival analysis** | | | | | | |
| | Univariable | | Multivariable* | | Multivariable*& | |
| **Variable** | **TR (95%CI)** | ***p*** | **TR (95%CI)** | ***p*** | **TR (95%CI)** | ***p*** |
| **Extent of axillary evaluation** | | | | | | |
| ≥10 | 1 | | 1 | | 1 | |

| | | | | | | | |
|---|---|---|---|---|---|---|---|
| | 2-3 | 0.548 (0.374 – 0.802) | 0.002 | 0.549 (0.394 – 0.764) | <0.0005 | 0.308 (0.161 – 0.590) | <0.0005 |
| **Extent of axillary evaluation* Surgery** |  |  |  |  |  |  | ----- |
| **Extent of axillary evaluation*Radiation therapy** | | | | | | | |
| | 2-3 analyzed LN*received radiation | | | | | 2.230 (1.044 – 4.763) | 0.038 |
| **Extent of axillary evaluation*T stage** |  |  |  |  |  |  | ----- |
| **Extent of axillary evaluation*Molecular subtype** |  |  |  |  |  |  | ----- |

*Adjusted by age (ordinal), race, median household income, rural-urban continuum, marital status, histological grade, molecular subtype, T stage, chemotherapy, surgical approach, radiotherapy and response to neoadjuvance.

&The overall effect of the interactions was assessed using the likelihood ratio test.

**Supplementary Table 20.** Association between the extent of axillary evaluation and overall specific survival, weighted by inverse propensity of treatment — subgroup of patients with two positive lymph nodes and treated with neoadjuvant systemic therapy (*n*=1,568)

| | | | | | | | |
|---|---|---|---|---|---|---|---|
| **Time-dependent Cox regression** | | | | | | | |
| | | **Univariable** | | **Multivariable*#** | | **Multivariable*#** | |
| **Variable** | | **HR (95%CI)** | ***p*** | **HR (95%CI)** | ***p*** | **HR (95%CI)** | ***p*** |
| **Extent of axillary evaluation** | | | | | | | |
| | ≥10 | 1 | | 1 | | 1 | |
| | 2-3 | 1.877 (1.371 – 2.570) | <0.0001 | 2.145 (1.573 – 2.926) | <0.0001 | 2.145 (1.573 – 2.926) | <0.0001 |
| **Extent of axillary evaluation* Surgery** |  |  |  |  |  |  | ----- |
| **Extent of axillary evaluation*Radiation therapy** |  |  |  |  |  |  | ----- |
| **Extent of axillary evaluation*Molecular subtype** |  |  |  |  |  |  | ----- |
| **Log-logistic survival analysis** | | | | | | | |
| | | **Univariable** | | **Multivariable*** | | **Multivariable*&** | |
| **Variable** | | **TR (95%CI)** | ***p*** | **TR (95%CI)** | ***p*** | **TR (95%CI)** | ***p*** |
| **Extent of axillary evaluation** | | | | | | | |
| | ≥10 | 1 | | 1 | | 1 | |
| | 2-3 | 0.549 (0.432 – 0.697) | <0.0001 | 0.567 (0.457 – 0.703) | <0.0001 | 0.567 (0.457 – 0.703) | <0.0001 |
| **Extent of axillary evaluation* Surgery** |  |  |  |  |  |  | ----- |
| **Extent of axillary evaluation*Radiation therapy** |  |  |  |  |  |  | ----- |
| **Extent of axillary evaluation*Molecular subtype** |  |  |  |  |  |  | ----- |

*Adjusted by age (ordinal), race, rural-urban continuum, marital status, histological grade, molecular subtype, T stage, chemotherapy, surgical approach, radiotherapy and response to neoadjuvance. Median household income was not included due to perfect collinearity.

[#]The model was corrected with stratification [strata()] of molecular subtype, radiotherapy and response to neoadjuvance.

[&]The overall effect of the interactions was assessed using the likelihood ratio test.

**Supplementary Table 21.** Association between the extent of axillary evaluation and breast cancer-specific survival — subgroup of patients with one positive lymph nodes and treated with neoadjuvant systemic therapy after matching (*n*=1,482)

| **Time-dependent Cox regression** | | | | | | | |
|---|---|---|---|---|---|---|---|
| | | **Univariable** | | **Multivariable[*#]** | | **Multivariable[*#]** | |
| **Variable** | | **HR (95%CI)** | ***p*** | **HR (95%CI)** | ***p*** | **HR (95%CI)** | ***p*** |
| **Extent of axillary evaluation** | | 1 | | 1 | | 1 | |
| | ≥10 | | | | | | |
| | 2-3 | 0.993 (0.727 – 1.355) | 0.964 | 1.023 (0.726 – 1.441) | 0.898 | 1.023 (0.726 – 1.441) | 0.898 |
| **Extent of axillary evaluation* Surgery** |  |  |  |  |  |  | ----- |
| **Extent of axillary evaluation*Radiation therapy** |  |  |  |  |  |  | ----- |
| **Extent of axillary evaluation*T stage** |  |  |  |  |  |  | ----- |
| **Extent of axillary evaluation*Molecular subtype** |  |  |  |  |  |  | ----- |
| **Log-logistic survival analysis** | | | | | | | |
| | | **Univariable** | | **Multivariable[*]** | | **Multivariable[*&]** | |
| **Variable** | | **TR (95%CI)** | ***p*** | **TR (95%CI)** | ***p*** | **TR (95%CI)** | ***p*** |
| **Extent of axillary evaluation** | | | | | | | |
| | ≥10 | 1 | | 1 | | 1 | |
| | 2-3 | 0.951 (0.726 – 1.246) | 0.718 | 0.997 (0.773 – 1.287) | 0.985 | 0.997 (0.773 – 1.287) | 0.985 |
| **Extent of axillary evaluation* Surgery** |  |  |  |  |  |  | ----- |
| **Extent of axillary evaluation*Radiation therapy** |  |  |  |  |  |  | ----- |
| **Extent of axillary evaluation*T stage** |  |  |  |  |  |  | ----- |
| **Extent of axillary evaluation*Molecular subtype** |  |  |  |  |  |  | ----- |

[*]Adjusted by age (ordinal), race, median household income, rural-urban continuum, marital status, histological grade, molecular subtype, T stage, chemotherapy, surgical approach and radiotherapy.

[#]The model was corrected with the stratification [strata()] of age (ordinal), histological grade, and molecular subtype.

[&]The overall effect of the interactions was assessed using the likelihood ratio test

**Supplementary Table 22.** Association between the extent of axillary evaluation and breast cancer-specific survival, weighted by inverse propensity of treatment — subgroup of patients with one positive lymph nodes and treated with neoadjuvant systemic therapy (*n*=2,957)

| Time-dependent Cox regression | | | | | | |
|---|---|---|---|---|---|---|
| | **Univariable** | | **Multivariable*#** | | **Multivariable*#** | |
| **Variable** | **HR (95%CI)** | ***p*** | **HR (95%CI)** | ***p*** | **HR (95%CI)** | ***p*** |
| **Extent of axillary evaluation** | | | | | | |
| ≥10 | 1 | | 1 | | 1 | |
| 2-3 | 1.073 (0.823 – 1.400) | 0.600 | 1.011 (0.779 – 1.313) | 0.932 | 1.011 (0.779 – 1.313) | 0.932 |
| **Extent of axillary evaluation* Surgery** |  |  |  |  |  | ----- |
| **Extent of axillary evaluation*Radiation therapy** |  |  |  |  |  | ----- |
| **Extent of axillary evaluation*T stage** |  |  |  |  |  | ----- |
| **Extent of axillary evaluation*Molecular subtype** |  |  |  |  |  | ----- |
| **Log-logistic survival analysis** | | | | | | |
| | **Univariable** | | **Multivariable*** | | **Multivariable*&** | |
| **Variable** | **TR (95%CI)** | ***p*** | **TR (95%CI)** | ***p*** | **TR (95%CI)** | ***p*** |
| **Extent of axillary evaluation** | | | | | | |
| ≥10 | 1 | | 1 | | 1 | |
| 2-3 | 0.901 (0.729 – 1.113) | 0.334 | 0.985 (0.807 – 1.202) | 0.881 | 0.985 (0.807 – 1.202) | 0.881 |
| **Extent of axillary evaluation* Surgery** |  |  |  |  |  | ----- |
| **Extent of axillary evaluation*Radiation therapy** |  |  |  |  |  | ----- |
| **Extent of axillary evaluation*T stage** |  |  |  |  |  | ----- |
| **Extent of axillary evaluation*Molecular subtype** |  |  |  |  |  | ----- |

*Adjusted by age (ordinal), race, rural-urban continuum, marital status, histological grade, molecular subtype, T stage, chemotherapy, surgical approach and radiotherapy. Median household income was not included due to perfect collinearity.

#The model was corrected with the stratification [strata()] of age (ordinal), histological grade, molecular subtype, and chemotherapy=.

&The overall effect of the interactions was assessed using the likelihood ratio test.

**Supplementary Table 23.** Association between the extent of axillary evaluation and breast cancer-specific survival — subgroup of patients with two positive lymph nodes and treated with neoadjuvant systemic therapy after matching (*n*=454)

| Cox regression | | | | | | | |
|---|---|---|---|---|---|---|---|
| | | **Univariable** | | **Multivariable*** | | **Multivariable*** | |
| **Variable** | | **HR (95%CI)** | ***p*** | **HR (95%CI)** | ***p*** | **HR (95%CI)** | ***p*** |
| **Extent of axillary evaluation** | | | | | | | |
| | ≥10 | 1 | | 1 | | 1 | |
| | 2-3 | 1.971 (1.208 – 3.043) | 0.006 | 2.137 (1.247 – 3.661) | 0.006 | 2.497 (0.359 – 17.395) | 0.355 |
| **Extent of axillary evaluation* Surgery** |  |  |  |  |  |  | ----- |
| **Extent of axillary evaluation*Radiation therapy** | | | | | | | 0.011 |
| 2-3 analyzed LN*received radiation | | | | | | 0.261 (0.077 – 0.888) | 0.032 |
| **Extent of axillary evaluation*T stage** |  |  |  |  |  |  | ----- |
| **Extent of axillary evaluation*Molecular subtype** |  |  |  |  |  |  | ----- |
| **Log-logistic survival analysis** | | | | | | | |
| | | **Univariable** | | **Multivariable*** | | **Multivariable*&** | |
| **Variable** | | **TR (95%CI)** | ***p*** | **TR (95%CI)** | ***p*** | **TR (95%CI)** | ***p*** |
| **Extent of axillary evaluation** | | | | | | | |
| | ≥10 | 1 | | 1 | | 1 | |
| | 2-3 | 0.523 (0.333 – 0.820) | 0.005 | 0.547 (0.375 – 0.796) | 0.0016 | 0.281 (0.131 – 0.602) | 0.0011 |
| **Extent of axillary evaluation* Surgery** |  |  |  |  |  |  | ----- |
| **Extent of axillary evaluation*Radiation therapy** | | | | | | | |
| 2-3 analyzed LN*received radiation | | | | | | 2.508 (1.035 – 6.078) | 0.042 |

| | |
|---|---|
| **Extent of axillary evaluation*T stage** | ----- |
| **Extent of axillary evaluation*Molecular subtype** | ----- |

*Adjusted by age (ordinal), race, median household income, rural-urban continuum, marital status, histological grade, molecular subtype, T stage, chemotherapy, surgical approach, radiotherapy and response to neoadjuvance.

#The model was corrected with the stratification [strata()] of age (ordinal).

&The overall effect of the interactions was assessed using the likelihood ratio test.

**Supplementary Table 24.** Association between the extent of axillary evaluation and breast cancer-specific survival, weighted by inverse propensity of treatment — subgroup of patients with two positive lymph nodes and treated with neoadjuvant systemic therapy (*n*=1,568)

| Time-dependent Cox regression | | | | | | | |
|---|---|---|---|---|---|---|---|
| | | Univariable | | Multivariable[*#] | | Multivariable[*#] | |
| Variable | | HR (95%CI) | *p* | HR (95%CI) | *p* | HR (95%CI) | *p* |
| **Extent of axillary evaluation** | | | | | | | |
| | ≥10 | 1 | | 1 | | 1 | |
| | 2-3 | 10.577 (2.035 – 54.980) | 0.005 | 1.976 (1.359 – 2.873) | <0.0005 | 1.976 (1.359 – 2.873) | <0.0005 |
| **tt* analyzed lymph nodes** | | | | | | | |
| | 2-3 | 0.616 (0.387 – 0.979) | 0.041 | | | | |
| **Extent of axillary evaluation* Surgery** |  |  |  |  |  |  | ----- |
| **Extent of axillary evaluation*Radiation therapy** |  |  |  |  |  |  | ----- |
| **Extent of axillary evaluation*Molecular subtype** |  |  |  |  |  |  | ----- |
| **Log-logistic survival analysis** | | | | | | | |
| | | Univariable | | Multivariable[*] | | Multivariable[*&] | |
| Variable | | TR (95%CI) | *p* | TR (95%CI) | *p* | TR (95%CI) | *p* |
| **Extent of axillary evaluation** | | | | | | | |
| | ≥10 | 1 | | 1 | | 1 | |
| | 2-3 | 0.517 (0.395 – 0.678) | <0.0001 | 0.532 (0.417 – 0.678) | <0.0001 | 0.532 (0.417 – 0.678) | <0.0001 |
| **Extent of axillary evaluation* Surgery** |  |  |  |  |  |  | ----- |
| **Extent of axillary evaluation*Radiation therapy** |  |  |  |  |  |  | ----- |
| **Extent of axillary evaluation*Molecular subtype** |  |  |  |  |  |  | ----- |

[*]Adjusted by age (ordinal), race, rural-urban continuum, marital status, histological grade, molecular subtype, T stage, chemotherapy, surgical approach, radiotherapy and response to neoadjuvance. Median household income was not included due to perfect collinearity.

[#]The model was corrected with stratification [strata()] of molecular subtype, radiotherapy and response to neoadjuvance.

[&]The overall effect of the interactions was assessed using the likelihood ratio test.

**Supplementary Figures**

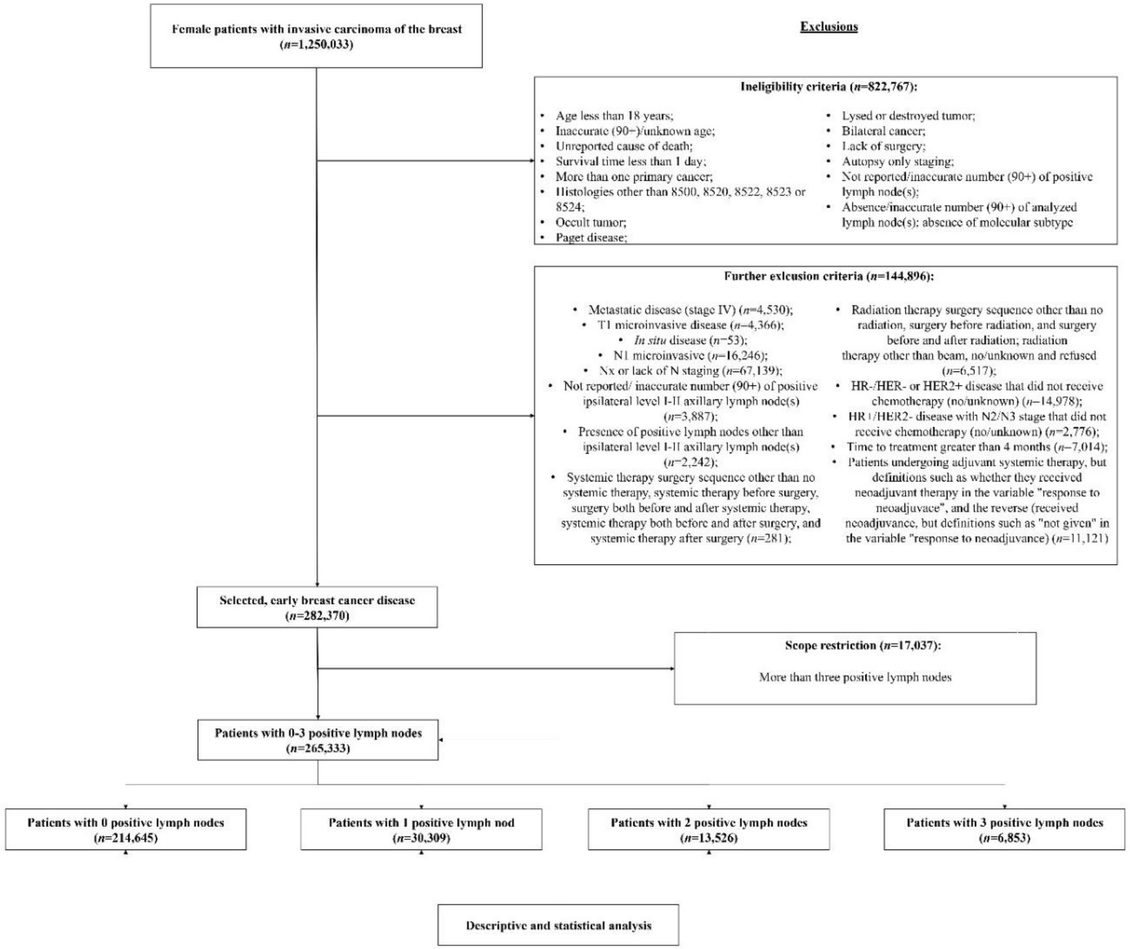


**Supplementary Figure 1. Flowchart of patient inclusion.**

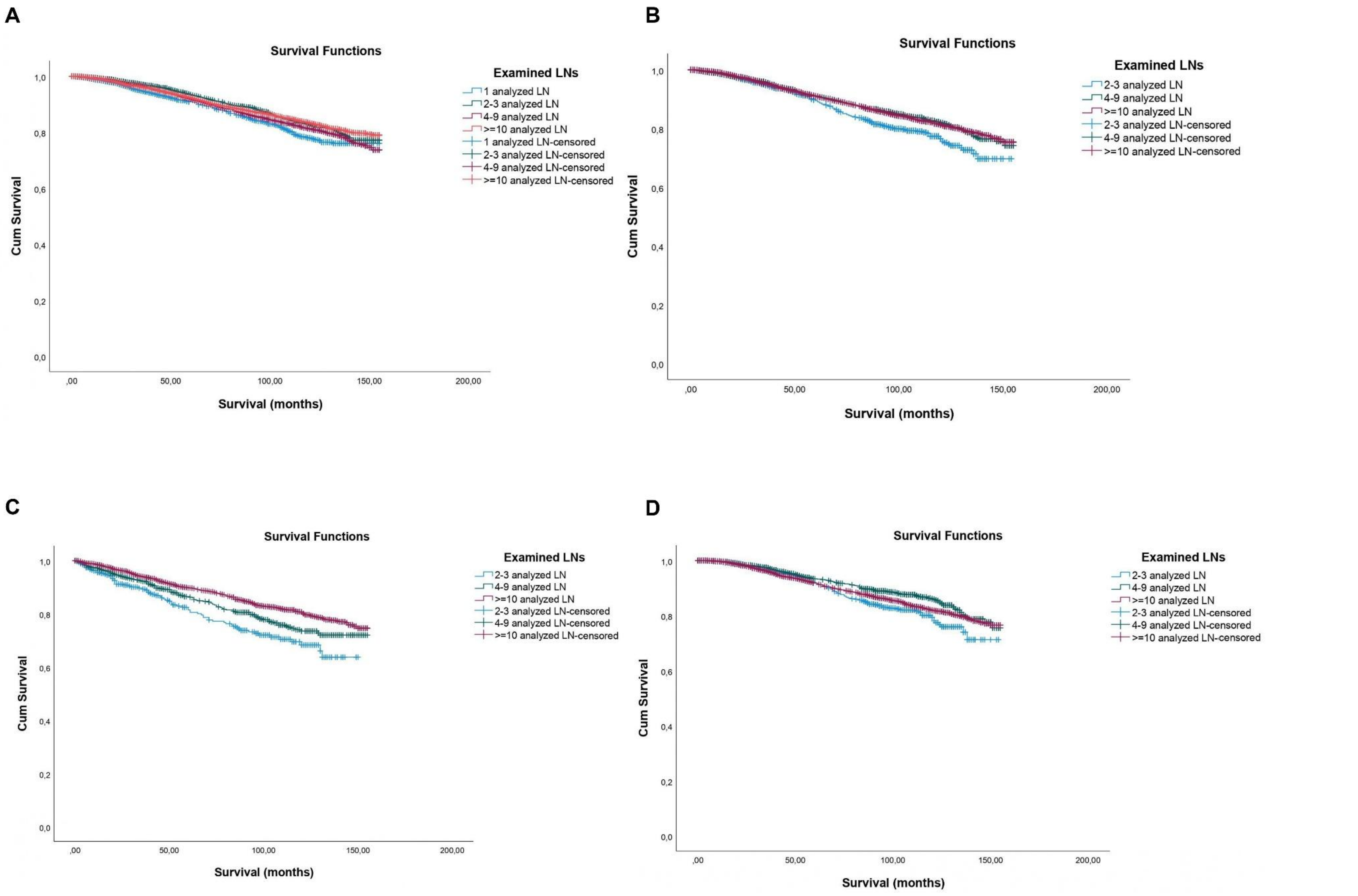
A
Survival Functions
Examined LNs
1 analyzed LN
2-3 analyzed LN
4-9 analyzed LN
>=10 analyzed LN
1 analyzed LN-censored
2-3 analyzed LN-censored
4-9 analyzed LN-censored
>=10 analyzed LN-censored
Cum Survival
1,0
0,8
0,6
0,4
0,2
0,0
,00
50,00
100,00
150,00
200,00
Survival (months)
B
Survival Functions
Examined LNs
2-3 analyzed LN
4-9 analyzed LN
>=10 analyzed LN
2-3 analyzed LN-censored
4-9 analyzed LN-censored
>=10 analyzed LN-censored
Cum Survival
1,0
0,8
0,6
0,4
0,2
0,0
,00
50,00
100,00
150,00
200,00
Survival (months)
C
Survival Functions
Examined LNs
2-3 analyzed LN
4-9 analyzed LN
>=10 analyzed LN
2-3 analyzed LN-censored
4-9 analyzed LN-censored
>=10 analyzed LN-censored
Cum Survival
1,0
0,8
0,6
0,4
0,2
0,0
,00
50,00
100,00
150,00
200,00
Survival (months)
D
Survival Functions
Examined LNs
2-3 analyzed LN
4-9 analyzed LN
>=10 analyzed LN
2-3 analyzed LN-censored
4-9 analyzed LN-censored
>=10 analyzed LN-censored
Cum Survival
1,0
0,8
0,6
0,4
0,2
0,0
,00
50,00
100,00
150,00
200,00
Survival (months)

**Supplementary Figure 2. Kaplan-Meier curves for overall survival according to the extent of axillary evaluation in patients treated with adjuvant systemic therapy.** (A) Patients with one positive lymph node and treated with adjuvant systemic therapy (*n*=22,469) (Log-Rank $\chi^2_{(df:3)}$: 24.20, p<0.0001; Breslow: $\chi^2_{(df:3)}$: 25.65, p<0.0001; Tarone-Ware: $\chi^2_{(df:3)}$: 25.39; p<0.0001). (B) Patients with two positive lymph node and treated with adjuvant systemic therapy (*n*=10,116) (Log-Rank $\chi^2_{(df:3)}$: 10.45, p=0.005; Breslow: $\chi^2_{(df:3)}$: 6.20, p=0.045; Tarone-Ware: $\chi^2_{(df:3)}$: 8.20; p=0.017). (C) Patients with two positive lymph node and radiation therapy not given/unknown if given (*n*=2,745) (Log-Rank $\chi^2_{(df:2)}$: 22.83, p<0.0001; Breslow: $\chi^2_{(df:2)}$: 24.64, p<0.0001; Tarone-Ware: $\chi^2_{(df:2)}$: 24.30; p<0.0001). (D) Patients with two positive lymph node that received radiation therapy (*n*=7,124) (Log-Rank $\chi^2_{(df:2)}$: 7.86, p=0.020; Breslow: $\chi^2_{(df:2)}$: 6.75, p=0.034; Tarone-Ware: $\chi^2_{(df:2)}$: 7.63; p=0.022). Legend: LN – lymph node.

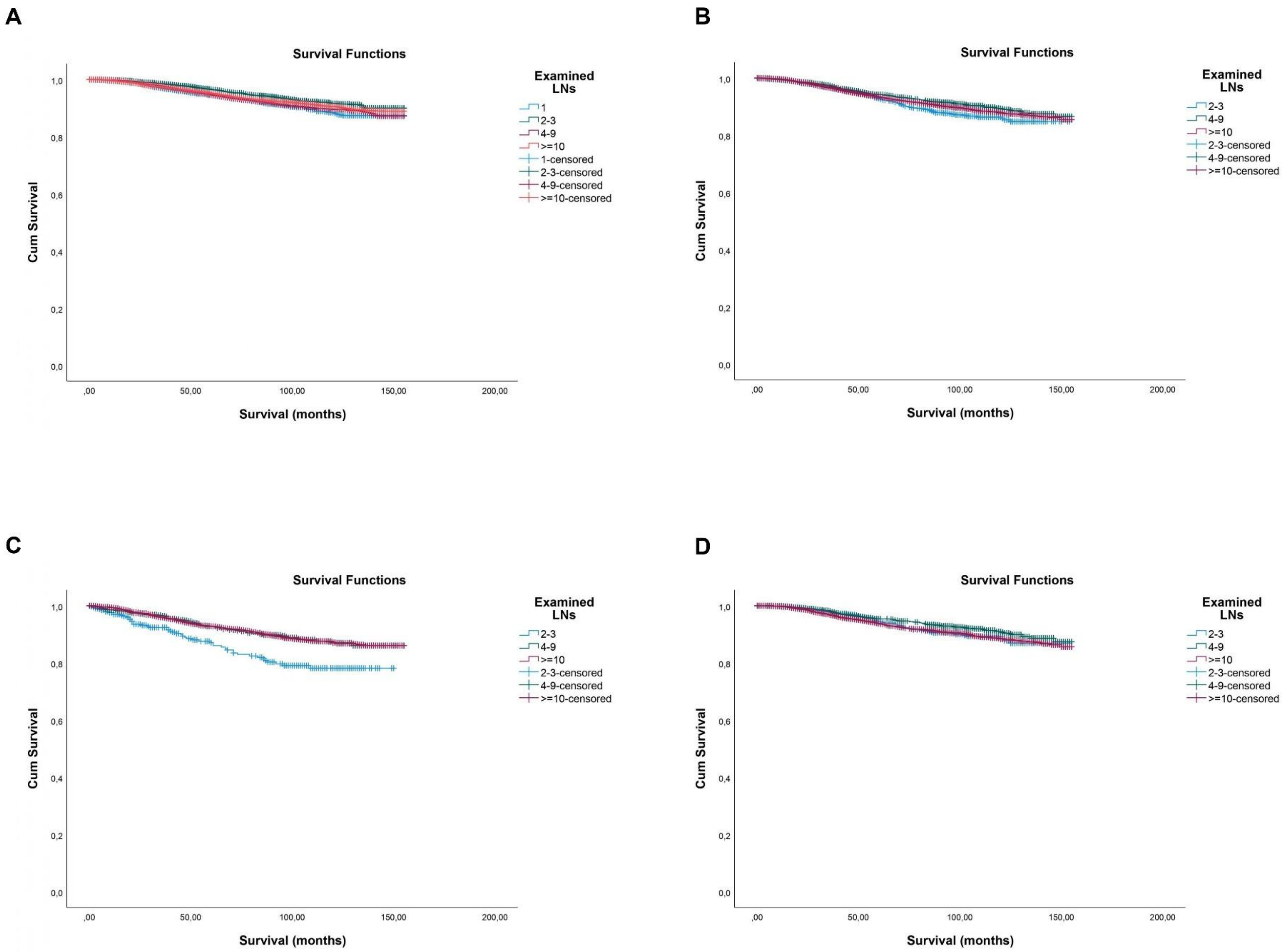
A
Survival Functions
Cum Survival
1,0
0,8
0,6
0,4
0,2
0,0
,00
50,00
100,00
150,00
200,00
Survival (months)
Examined LNs
1
2-3
4-9
>=10
1-censored
2-3-censored
4-9-censored
>=10-censored
B
Survival Functions
Cum Survival
Survival (months)
Examined LNs
2-3
4-9
>=10
2-3-censored
4-9-censored
>=10-censored
C
Survival Functions
Cum Survival
Survival (months)
Examined LNs
2-3
4-9
>=10
2-3-censored
4-9-censored
>=10-censored
D
Survival Functions
Cum Survival
Survival (months)
Examined LNs
2-3
4-9
>=10
2-3-censored
4-9-censored
>=10-censored

**Supplementary Figure 3. Kaplan-Meier curves for breast cancer-specific survival according to the extent of axillary evaluation in patients treated with adjuvant systemic therapy.** (A) Patients with one positive lymph node and treated with adjuvant systemic therapy (*n*=22,469) (Log-Rank $\chi^2_{(df:3)}$: 18.96, p<0.0001; Breslow: $\chi^2_{(df:3)}$: 22.15, p<0.0001; Tarone-Ware: $\chi^2_{(df:3)}$: 21.09; p<0.0001). (B) Patients with two positive lymph node and treated with adjuvant systemic therapy (*n*=10,116) (Log-Rank $\chi^2_{(df:3)}$: 5.33, p=0.070; Breslow: $\chi^2_{(df:3)}$: 4.35, p=0.114; Tarone-Ware: $\chi^2_{(df:3)}$: 5.00; p=0.017). (C) Patients with two positive lymph node and radiation therapy not given/unknown if given (*n*=2,745) (Log-Rank $\chi^2_{(df:2)}$: 17.69, p<0.0001; Breslow: $\chi^2_{(df:2)}$: 20.03, p<0.0001; Tarone-Ware: $\chi^2_{(df:2)}$: 19.24; p<0.0001). (D) Patients with two positive lymph node that received radiation therapy (*n*=7,124) (Log-Rank $\chi^2_{(df:2)}$: 4.53, p=0.104; Breslow: $\chi^2_{(df:2)}$: 6.21, p=0.045; Tarone-Ware: $\chi^2_{(df:2)}$: 5.58; p=0.061). Legend: LN – lymph node.
